# Epigenetic scores indicate differences in the proteome of preterm infants

**DOI:** 10.1101/2023.12.19.23300227

**Authors:** Katie Mckinnon, Eleanor L.S. Conole, Kadi Vaher, Robert F. Hillary, Danni A. Gadd, Justyna Binkowska, Gemma Sullivan, Anna J. Stevenson, Amy Corrigan, Lee Murphy, Heather C. Whalley, Hilary Richardson, Riccardo E. Marioni, Simon R. Cox, James P. Boardman

## Abstract

**Background:** Epigenetic scores (EpiScores), reflecting DNA methylation (DNAm)-based surrogates for complex traits, have been developed for multiple circulating proteins. EpiScores for pro-inflammatory proteins, such as C-reactive protein (DNAm CRP), are associated with brain health and cognition in adults and with inflammatory comorbidities of preterm birth in neonates. Social disadvantage can become embedded in child development through inflammation, and deprivation is over-represented in preterm infants. We tested the hypotheses that preterm birth and socioeconomic status (SES) are associated with alterations in a set of EpiScores enriched for inflammation-associated proteins.

**Results:** 104 protein EpiScores were derived from saliva samples of 332 neonates born at gestational age (GA) 22.14 to 42.14 weeks. Saliva sampling was between 36.57 and 47.14 weeks. Forty-three (41%) EpiScores were associated with low GA at birth (standardised estimates |0.14 to 0.88|, Bonferroni-adjusted *p*-value <8.3×10^−3^). These included EpiScores for chemokines, growth factors, proteins involved in neurogenesis and vascular development, cell membrane proteins and receptors, and other immune proteins. Three EpiScores were associated with SES, or the interaction between birth GA and SES: afamin, intercellular adhesion molecule 5 and hepatocyte growth factor-like protein (standardised estimates |0.06 to 0.13|, Bonferroni-adjusted *p*-value <8.3×10^−3^). In a preterm sub-group (n=217, median [range] GA 29.29 weeks [22.14 to 33.0 weeks]), SES-EpiScore associations did not remain statistically significant after adjustment for sepsis, bronchopulmonary dysplasia, necrotising enterocolitis, and histological chorioamnionitis.

**Conclusions:** Low birth GA is substantially associated with a set of EpiScores. The set was enriched for inflammatory proteins, providing new insights into immune dysregulation in preterm infants. SES had fewer associations with EpiScores; these tended to have small effect sizes and were not statistically significant after adjusting for inflammatory comorbidities. This suggests that inflammation is unlikely to be the primary axis through which SES becomes embedded in the development of preterm infants in the neonatal period.

## 1.0 Background

Preterm birth (delivery at less than 37 weeks of gestation) affects around 10% of births worldwide and is closely associated with increased likelihood of cerebral palsy, neurocognitive impairment, behavioural, social and communication difficulties, and mental and cardiometabolic health diagnoses across the life course [1–5]. These adverse outcomes can be explained, in part, by deleterious effects of early exposure to extrauterine life on brain and cardiac development, and they are often accompanied by changes in blood proteins, including those reflecting the perinatal innate and adaptive immune response [6–8].

Socioeconomic status (SES) is also associated with the adverse neurodevelopmental and health outcomes listed above [9–12], and social deprivation is consistently over-represented among preterm children and their families [13,14]. In a meta-analysis of 43 studies (n=111,156 individuals), low SES associated with increased inflammatory markers of disease risk (C-reactive protein [CRP] and interleukin-6 [IL6]), which suggests that pro-inflammatory pathways may be important mechanisms for translating social inequalities into health disparities [15]. However, only four studies included participants under 10 years of age, leaving uncertainty about SES-inflammation correlations in early life [16–19].

Although protein levels are commonly used as biomarkers of exposure and disease risk, they are limited because they are often phasic in the systemic circulation, rely on venepuncture, and may not capture baseline status or chronicity. For example, inflammation is often measured using acute-phase inflammatory proteins such as CRP [20,21], but it is not always reliable [22], particularly in neonates, and a single-time-point measure may not reflect baseline inflammation or capture chronic inflammation [23]. These challenges have been addressed by the development of DNA methylation (DNAm) markers of protein expression (EpiScores), which are derived from a linear weighted sum of DNAm sites that are correlated with protein levels. Several EpiScores are associated with magnetic resonance imaging (MRI) measures of brain health, cognition, child mental health, stroke, ischaemic heart disease Alzheimer’s disease, lung cancer [24–31]. In neonates, DNAm CRP is associated with birth gestational age (GA), perinatal inflammatory processes, and MRI features of encephalopathy of prematurity [32]. Childhood SES is associated with differential DNAm in inflammation-related genes [33,34] and at CpG sites that correlate with an inflammation index [35]. Adult SES and social mobility are associated with variations in DNAm in inflammation-related genes [33,36]. Importantly, SES-related DNAm variations are associated with differences in gene expression so may have functional consequences [33,36].

We investigated relationships between preterm birth, SES, and 104 EpiScores enriched for inflammation-related proteins [26–28,31,37]. We tested the following hypotheses: first, low GA is associated with differences in EpiScores; second, SES is correlated with EpiScores, and interacts with birth GA, but the relationship is attenuated by inflammatory disease burden in preterm infants.

## 2.0 Methods

### 2.1 Participants

Participants were preterm infants (born ≤33 weeks’ gestation) and term-born infants born at the Royal Infirmary of Edinburgh, UK. These infants were recruited to a longitudinal cohort study designed to investigate the effect of preterm birth on brain development and outcomes with multimodal data collection [38]. Infants were recruited between February 2012 and December 2021.

Exclusion criteria were congenital malformation, chromosomal abnormality, congenital infection, cystic periventricular leukomalacia, haemorrhagic parenchymal infarction, and post-haemorrhagic ventricular dilatation. These criteria mean the cohort is representative of the majority of survivors of modern intensive care practices [38].

Final participants included were 217 preterm infants (born ≤33 weeks’ GA) and 115 term-born infants, with median birth GA of 29.29 weeks and 39.71 weeks respectively. Their demographic characteristics are shown in Table 1. The three SES measures (Scottish Index of Multiple Deprivation (SIMD 2016) [39], maternal education and maternal occupation) differed between the preterm and term groups (Cohen’s d effect sizes 0.52-0.68). Ethnicity did not differ between groups and is representative of the Edinburgh area [40].

**Table 1.** Participant characteristics.

| Demographic measure |  | Preterm (n=217) | Term (n=115) |
| --- | --- | --- | --- |
| Sex | Male | 114/217 (52.5%) | 64/115 (55.7%) |
|  | Female | 103/217 (47.5%) | 51/115 (44.3%) |
| Birth GA (weeks) - median (range) |  | 29.29 (22.14-33.0) | 39.71 (37.0-42.14) |
| Birthweight (g) - median (range) |  | 1200 (370-2510) | 3450 (2346-4670) |
| Birthweight z-score - median (range) |  | 0.10 (-3.13-2.07) | 0.43 (-2.3-2.96) |
| SIMD rank - median (range)<sup>A</sup> |  | 3720 (6-6966) | 5344 (267-6967) |
| Maternal ethnicity | African | 1/217 (0.5%) | 0/115 (0%) |
|  | Bangladeshi | 0/217 (0%) | 1/115 (0.9%) |
|  | Caribbean | 0/217 (0%) | 0/115 (0%) |
|  | Chinese | 0/217 (0%) | 1/115 (0.9%) |
|  | Indian | 3/217 (2.4%) | 1/115 (0.9%) |
|  | Pakistani | 4/214 (1.8%) | 1/115 (0.9%) |
|  | White | 195/217 (89.9%) | 106/115 (92.2%) |
|  | White/Asian | 2/217 (0.9%) | 0/115 (0%) |
|  | White/Black African | 0/217 (0%) | 1/115 (0.9%) |
|  | White/Black Caribbean | 1/217 (0.5%) | 1/115 (0.9%) |
|  | Other Asian<sup>B</sup> | 1/217 (0.5%) | 2/115 (1.7%) |
|  | Other ethnic group<sup>C</sup> | 6/217 (2.8%) | 0/115 (0%) |
|  | Other mixed ethnic background<sup>D</sup> | 4/217 (1.8%) | 1/115 (0.9%) |
| Maternal education | None | 7/208 (3.4%) | 0/115 (0%) |
|  | Basic high school qualification (1-4) | 5/208 (2.4%) | 2/115 (1.7%) |
|  | Basic high school qualification (>=5) | 8/208 (3.8%) | 1/115 (0.9%) |
|  | Advanced high school qualification | 32/208 (15.4%) | 3/115 (2.6%) |
|  | College qualification | 46/208 (22.1%) | 8/115 (7.0%) |
|  | University undergraduate | 61/208 (29.3%) | 50/115 (43.5%) |
|  | University postgraduate | 49/208 (23.6%) | 51/115 (44.3%) |
| Maternal occupation | Unemployed | 12/213 (5.6%) | 1/115 (0.9%) |
|  | Homemaker | 10/213 (4.7%) | 1/115 (0.9%) |
|  | Still in full time education | 9/213 (4.2%) | 1/115 (0.9%) |
|  | Sheltered employment | 1/213 (0.5%) | 0/115 (0%) |
|  | Unskilled | 11/213 (5.2%) | 3/115 (2.6%) |
|  | Partly skilled | 7/213 (3.3%) | 2/115 (1.7%) |
|  | Manual skilled | 25/213 (11.7%) | 9/115 (7.8%) |
|  | Non-manual skilled | 44/213 (20.7%) | 11/115 (9.6%) |
|  | Professional | 94/213 (44.1%) | 87/115 (75.7%) |
| Sample GA (weeks) - median (range) |  | 40.57 (36.57-45.86) | 42.0 (39.86-47.14) |
| Batch | 1 | 92/217 (42.4%) | 46/115 (40.0%) |
|  | 2 | 62/217 (28.6%) | 56/115 (48.7%) |
|  | 3 | 32/217 (14.7%) | 1/115 (0.9%) |
|  | 4 | 31/217 (14.3%) | 12/115 (10.4%) |

A: Preterm n=216, term n=115
B: "Other Asian" includes Sri Lankan (n=1), Malay (n=1)
C: "Other ethnic group" includes Arab (n=1), Iraqi (n=1), white Bulgarian (n=1), Fijian (n=1), Japanese (n=1), Hong Kong (n=1).
D: "Other mixed ethnic background" includes Pakistani/Scottish (n=2), British/Arab (n=1), Sri Lankan/Black (n=1), Sri Lankan/Indian (n=1).
GA = gestational age, SIMD = Scottish index of multiple deprivation.

### 2.2 DNA methylation

Saliva samples for DNAm were collected at term-equivalent age using Oragene OG-575 Assisted Collection kits (DNA Genotek, ON, Canada) and DNA was extracted using prepIT.L2P reagent (DNA Genotek, ON, Canada). Saliva sampling was used due to accessibility and the non-invasiveness of the method; DNAm patterns measured via saliva samples correlate with brain and other tissue DNAm patterns [41,42]. We chose to sample at the term-corrected gestation timepoint to include the allostatic load of both prenatal and early postnatal exposures.

DNA was bisulphite converted and methylation levels were measured using Illumina HumanMethylationEPIC BeadChip (Illumina, San Diego, CA, USA) at the Edinburgh Clinical Research Facility (Edinburgh, UK). The arrays were imaged on the Illumina iScan or HiScan platform, and genotypes were called automatically using GenomeStudio Analysis software version 2011.1 (Illumina). DNAm was processed in four batches.

Raw intensity (.idat) files were read into the R environment using minfi. wateRmelon and minfi were used for preprocessing, quality control and normalisation [43]. The pfilter function in wateRmelon was used to exclude samples with 1% of sites with a detection *p*-value >0.05, sites with beadcount <3 in 5% of samples, and sites with 1% of samples with detection *p*-value >0.05. Cross hybridising probes, probes targeting single nucleotide polymorphisms with overall minor allele frequency ≥0.05, and control probes were also removed. Samples were removed if there was a mismatch between predicted sex (minfi) and recorded sex (n=3), or if samples did not meet preprocessing quality control criteria (n=29). Data were danet normalised, which includes background correction and dye bias correction [43]. Saliva contains different cells types, including buccal epithelial cells. Epithelial cell proportions were estimated with epigenetic dissection of intra-sample heterogeneity with the reduced partial correlation method implemented in the R package EpiDISH [44]. Probes located on sex chromosomes were removed before analysis. The cohort includes twins (n=32); these were randomly removed leaving one participant per twin pair. This left a final sample size of n=332.

### 2.3 EpiScore calculation

The 104 protein EpiScores included 100 EpiScores from Gadd *et al* [26], a study enriched for inflammatory-related proteins, excluding those where the required CpGs were not available, owing to differences in assay platform CpG coverage relative to Gadd *et al* [26]. For duplicate proteins, those developed using Olink platform-identified proteins (antibody-based assays) were prioritised over those from SOMAscan platforms (aptamer-based assays), due to specificity and reproducibility, and the variable correlation between the two methods [45–48] (for details see Supplementary eMethods, Additional File 1). In addition, we included EpiScores for growth and differentiation factor 15 (GDF15) and N-terminal-pro B-type natriuretic peptide (NTproBNP) [37], and CRP. The CRP EpiScore used was Barker *et al*’s seven-CpG variation of Ligthart *et al*’s CRP EpiScore [27,31], as this is known to correlate with birth GA, perinatal pro-inflammatory exposures, and neonatal brain development [32].

For each individual, EpiScores were obtained by multiplying the methylation proportion at a given CpG by the effect size from previous studies. This was performed using the MethylDetectR platform [49] for those inflammatory proteins currently included and using R for those not currently included (CRP, GDF15, IL6, NTproBNP). All CpG sites and coefficients required to calculate the 104 EpiScores are in Supplementary Table 1 (Additional File 2).

### 2.4 Statistics

The predictor variables were SES and birth GA. SES was operationalised in three ways: neighbourhood-level SES using the Scottish Index of Multiple Deprivation (SIMD) [39], and two measures of family-level SES, which were maternal education (highest educational qualification) and maternal occupation (current or most recent occupation). For further details, see Supplementary eMethods, Additional File 1. Birth GA was a continuous variable to maximise statistical power [50,51]. We adjusted for GA at saliva sampling, DNAm batch, infant sex, and birthweight z-score.

All statistical analyses were performed in R (version 4.3.1) and were preregistered [52].

Principal component analysis (PCA) was used to determine the significance threshold for controlling type 1 error in analyses of multiple EpiScores [53]. We began with a correlation analysis, which showed correlation coefficients between EpiScores of |0.01 to 0.93| (Figure 1A). To determine the number of statistical “families” among the 104 EpiScores, PCA was performed. This yielded two principal components with eigenvalues >1, our pre-specified threshold, which explained 59.5% and 17.2% of variance, respectively (Figure 1B). Standardised component loadings are provided in Supplementary Table 3 (Additional File 1). In all subsequent analyses, we corrected for multiple comparisons across EpiScores and SES measures using a Bonferroni-adjusted *p*-value threshold of 8.3×10^−3^. This is 0.05/(2 x 3), with two reflecting the two principal components for Episcores and three reflecting the number of SES measures used.

**Figure 1.**
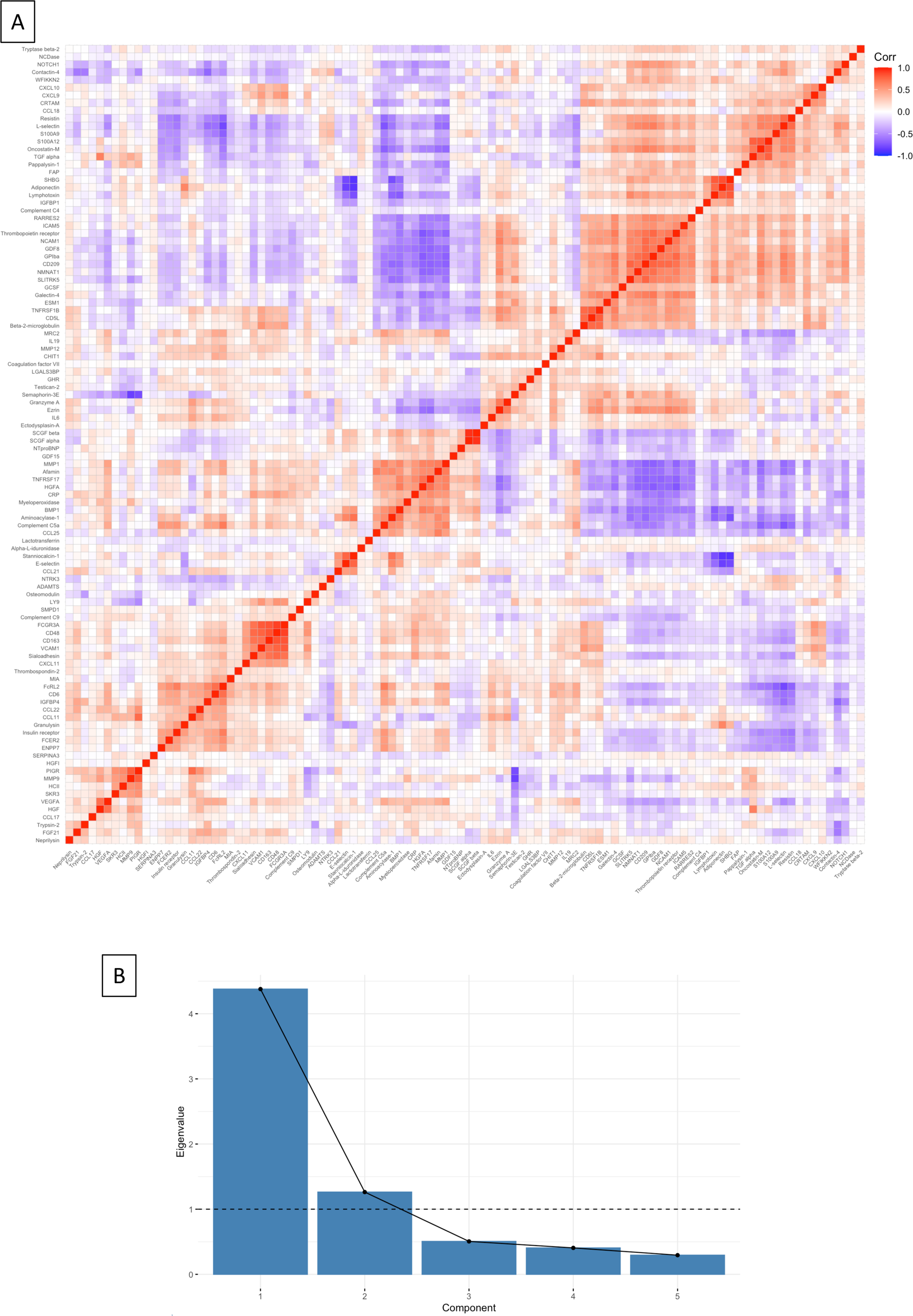
Determining significance threshold. Principal component analysis was used to determine the adjusted statistical significance threshold, given multiple statistical comparisons. Figure 1A shows a correlation matrix of 104 EpiScores, showing correlation coefficient as red for positive and blue negative associations when significant (*p*<0.05). Figure 1B shows a scree plot of principal components, with the eigenvalues for each component. Standardised component loadings for principal components one and two are provided in Supplementary Table 3 (Additional File 1).

We constructed generalised linear regression models for each EpiScore as outcome measure to assess associations between GA, each of the three SES measures (separate models for each of SIMD, maternal education, and maternal occupation), and the product interaction term SES*birth GA (removing the term if not significant), and adjusting for GA at sampling, sex, and batch.

For the preterm sub-group, we then additionally adjusted for perinatal inflammatory exposures known to be associated with the CRP EpiScore as, to our knowledge, this is the only DNAm proxy of an inflammatory protein that has been studied in this context [32]. These were histological chorioamnionitis (HCA), sepsis, bronchopulmonary dysplasia (BPD) and necrotising enterocolitis (NEC). For definitions see Supplementary eMethods (Additional File 1) and for frequencies see Supplementary Table 2 (Additional File 1).

## 3.0 Results

### 3.1 Associations between gestational age and EpiScores

Gestational age associated with 43 of the 104 EpiScores after adjustment for SIMD, maternal education, or maternal occupation (Figure 2A-C). The proteins represented by the 43 EpiScores are listed in Table 2 categorised by functional annotation adapted from the STRING database[54], and their broader roles in immune processes and inflammation, and the pathogenesis of neonatal diseases, where known, are described in Supplementary Table 4 (Additional File 1). 29 (67%) EpiScores negatively associated with birth GA (standardised estimates |0.14-0.76|, adjusted *p*-value <8.3×10^−3^), and 14 (33%) EpiScores positively associated with birth GA (standardised estimates 0.14-0.88, adjusted *p*-value <8.3×10^−3^).

**Figure 2.**
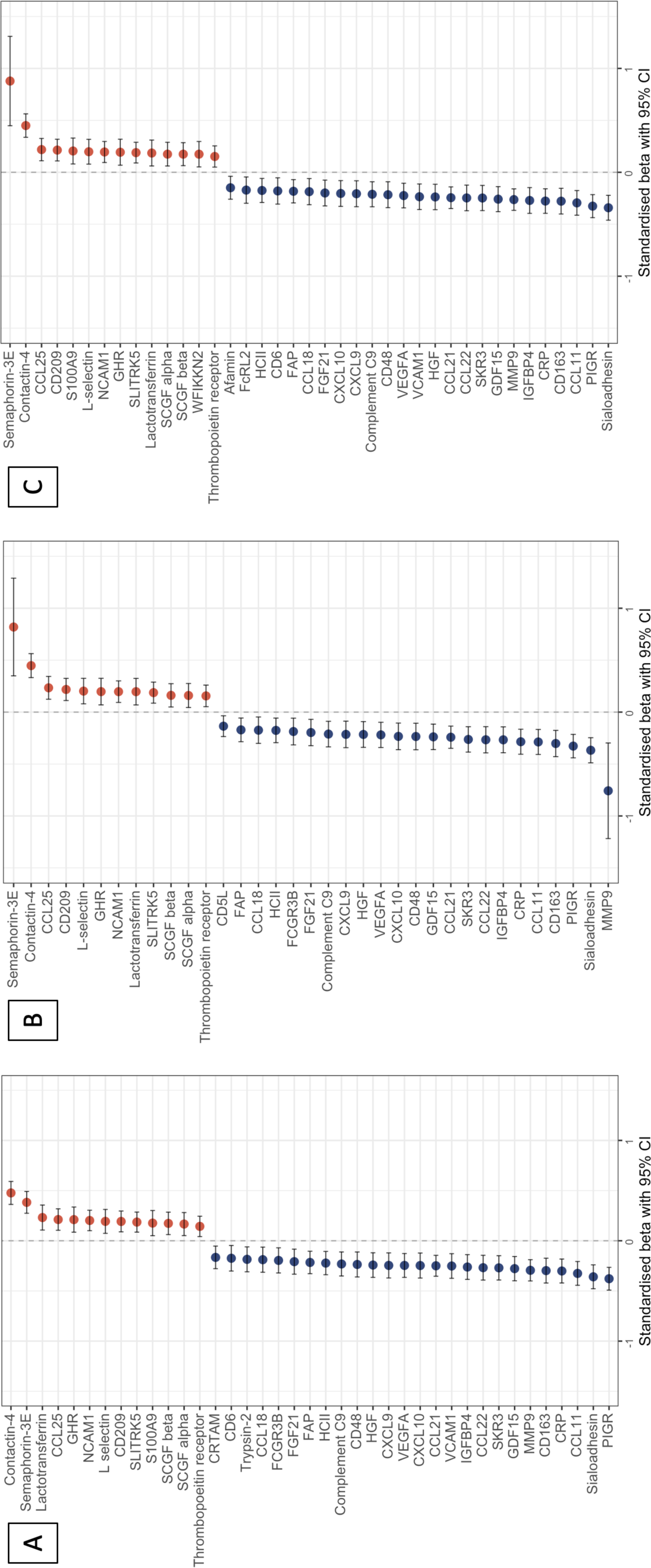
EpiScores associated with gestational age in regression models adjusted for socioeconomic status. EpiScores associated with gestational age in regression models with (A) Scottish Index of Multiple Deprivation, (B) maternal education, and (C) maternal occupation. Figure 2A (n=331) shows 39 associations, Figure 2B (n=323) shows 35 associations, and Figure 2C shows (n=328) shows 39 associations. Points and bars represent standardised beta and 95% confidence intervals, with red indicating positive and blue negative associations. Covariates included in all models: age at sample, birthweight z-score, sex, and methylation processing batch. Bonferroni-adjusted *p*-value <8.3×10^−3^. CCL11 – C-C chemokine 11, CCL18 – C-C chemokine 18, CCL21 – C-C chemokine 21, CCL22 – C-C chemokine 22, CCL25 – C-C chemokine 25, CD5L – CD5 antigen-like protein, CD6 – T-cell differentiation antigen, CD163 – scavenger receptor cysteine-rich type 1 protein M130, CI – confidence interval, CRP – C-reactive protein, CRTAM – Cytotoxic and regulatory T-cell molecule, CXCL9 – C-X-C motif chemokine 9, CXCL10 – C-X-C motif chemokine 10, FAP – Fibroblast Activation Protein alpha, FCGR3B – Low affinity immunoglobulin gamma Fc region receptor III-B, FcRL2 – Fc receptor-like protein 2, FGF21 – Fibroblast growth factor 21, GDF15 – Growth/differentiation factor 15, GHR – Growth hormone receptor, HCII – Heparin cofactor II, HGF – Hepatocyte growth factor alpha chain, IGFBP4 – Insulin-like growth factor-binding protein 4, MMP9 – Matrix metalloproteinase-9, NCAM1 – Neural cell adhesion molecule 1, PIGR – Polymeric immunoglobulin receptor, SCGF – stem cell growth factor, SIMD – Scottish Index of Multiple Deprivation, SKR3 – Serine/threonine-protein kinase receptor R3, SLITRK5 – SLIT and NTRK-like protein 5, VCAM1 – Vascular cell adhesion protein 1, VEGFA – Vascular endothelial growth factor A, WFIKKN2 – WAP Kazal immunoglobulin Kunitz and NTR domain-containing protein 2.

**Table 2.** Protein EpiScores associated with birth gestational age.

| Chemokines | Growth factors | Neurogenesis | Vascular development | Cell membrane proteins / receptors | Other immune response |
| --- | --- | --- | --- | --- | --- |
| CCL11 | FGF21 | NCAM1 | SKR3 | Afamin | Complement C9 |
| CCL18 | GDF15 | Semaphorin 3E | VCAM1 | CD5L | CRP |
| CCL21 | GHR | SLITRK5 | VEGFA | CD6 | CRTAM |
| CCL22 | HGF |  |  | CD48 | FAP |
| CCL25 | IGFBP4 |  |  | CD163 | HCII |
| CXCL9 | SCGF alpha |  |  | CD209 | L-selectin |
| CXCL10 | SCGF beta |  |  | Contactin-4 | Lactotransferrin |
|  | WFIKKN2 |  |  | FcRL2 | MMP9 |
|  |  |  |  | FcGR3B | S100A9 |
|  |  |  |  | PIGR | Sialoadhesin |
|  |  |  |  | Thrombopoietin receptor | Trypsin-2 |

2 43 EpiScores associated with birth gestational age in regression models adjusted for socioeconomic status. Roles adapted from the STRING database[54].
5 CCL11 – C-C chemokine 11, CCL18 – C-C chemokine 18, CCL21 – C-C chemokine 21, CCL22 – C-C chemokine 22, CCL25 – C-C chemokine 25, CD5L – 6 CD5 antigen-like protein, CD6 – T-cell differentiation antigen, CD163 – scavenger receptor cysteine-rich type 1 protein M130, CRP – C-reactive protein, 7 CRTAM – Cytotoxic and regulatory T-cell molecule, CXCL9 – C-X-C motif chemokine 9, CXCL10 – C-X-C motif chemokine 10, FAP – Fibroblast Activation 8 Protein alpha, FCGR3B – Low affinity immunoglobulin gamma Fc region receptor III-B, FcRL2 – Fc receptor-like protein 2, FGF21 – Fibroblast growth factor

33 EpiScores associated with low GA irrespective of SES measure used in the model. The results for all 104 EpiScores are provided in Supplementary Figures 1-9 (Additional File 1).

### 3.2 Associations between SES, EpiScores, and the effect of inflammatory comorbidities of preterm birth

Three out of 104 EpiScores associated with SES measures or the interaction between SES and GA (Figure 3). There was a small effect size association between higher afamin EpiScore and higher maternal occupation (standardised *β*=0.06, 95% confidence interval (CI) 0.02-0.11, *p*=0.0082), and DNAm afamin associated with the birth GA*maternal education interaction term such that afamin positively correlated with birth GA among babies with mothers without university education, and negatively correlated with birth GA among babies with mothers with university education (undergraduate or postgraduate) (standardised *β*=-0.12, 95% CI −0.20 to −0.04, *p*=0.0041, Supplementary Figure 10, Additional File 1).

**Figure 3.**
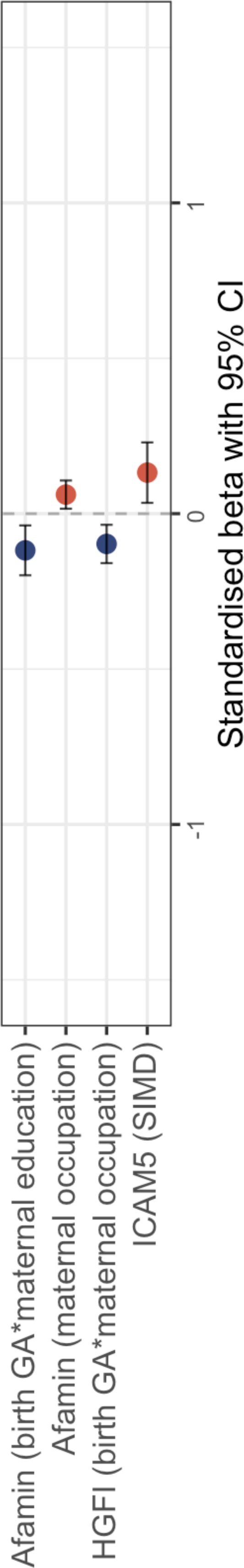
EpiScores associated with socioeconomic status or an interaction between socioeconomic status and birth gestational age. EpiScores associated with socioeconomic status (Scottish Index of Multiple Deprivation, maternal education, or maternal occupation), or with an interaction between socioeconomic status and birth gestational age. Regression models with gestational age at birth, gestational age at sample, birthweight z-score, sex, and methylation processing batch. Sample sizes: for the Scottish Index of Multiple Deprivation n=331, for maternal education n=323, and for maternal occupation n=328. 3/104 EpiScores were significant (Bonferroni-adjusted *p*-value <8.3×10^−3^). Points and bars represent standardised beta and 95% confidence intervals, with red indicating positive and blue negative associations. CI – confidence interval, GA – gestational age, HGFI – Hepatocyte growth factor-like protein alpha chain, ICAM5 – Intercellular adhesion molecule 5, SIMD – Scottish Index of Multiple Deprivation.

Higher intercellular adhesion molecule 5 (ICAM5) EpiScore associated with higher SIMD (standardised *β*=0.13, 95% CI 0.03-0.23, *p*=0.0079). Hepatocyte growth factor-like protein (HGFI) associated with the birth GA*maternal occupation interaction term (standardised *β*=-0.10, 95% CI - 0.16- -0.04, *p*=0.0021, Supplementary Figure 11, Additional File 1), such that HGFI EpiScore positively correlated with birth GA among babies with mothers who were unemployed, homemakers, in full time education, or in unskilled or manual occupations, but negatively correlated with birth GA among babies with mothers in partly skilled, non-manual skilled or professional occupations.

In the planned analysis of preterm-born babies only, when controlling for inflammatory exposures (sepsis, HCA, NEC, and BPD), a greater proportion of R^2^ was explained (R^2^=0.053-0.28 in unadjusted models, R^2^=0.029-0.437 in adjusted models), but the EpiScores no longer met our statistical threshold (*p*-values >0.045 with adjusted *p*-value threshold<8.3×10^−3^, see Supplementary Table 5, Additional File 1).

## 4.0 Discussion

In this study, we identified several associations between a set of EpiScores enriched for inflammatory proteins and low GA at birth. Few EpiScores associated with SES within the whole sample and these associations were partially attenuated in preterm infants who experienced inflammatory comorbidities. This is the first study to assess the impact of preterm birth and social status using epigenetic signatures designed to reflect the circulating proteome.

### 4.1 Associations between birth GA and EpiScores

43 EpiScores associated with preterm birth when sampled at term equivalent age. The EpiScores reflect chemokines, growth factors, proteins required for neurogenesis and vascular development, cell membrane proteins and receptors, and immune response proteins (Table 2). As well as having specific immunoregulatory roles, in the neonatal setting or relevant animal models, these proteins are associated with several comorbidities and developmental consequences of preterm birth. These include lung development and disease such as BPD [55–67], in utero and postnatal growth failure [68–71], HCA [7,72,73], patent ductus arteriosus [74–77], retinopathy of prematurity [78–82], NEC [83–85], hyperglycaemia [86], sepsis [85,87–90], brain injury [32,91–94], and neurodevelopmental outcomes [95–99].

### 4.2 Associations between SES measures and EpiScores

SES appears to play a much smaller role in the patterning of EpiScores compared to birth GA. SES measures, or interactions between birth GA and SES measures, were associated with only three of the 104 EpiScores studied: afamin, ICAM5 and HGFI. Afamin and ICAM5 positively associated with maternal occupation and SIMD, respectively. Afamin and HGFI both associated with the interaction term between SES and birth GA. Afamin is involved in vitamin E transport [100], ICAM5 has a role in microglial regulation [101], and HGFI is a macrophage-stimulating protein [102]. The relationships between these proteins and SES have not previously been investigated, although afamin is associated with the development of metabolic syndrome [103], which varies with SES [104].

Among preterm infants, no SES-EpiScore associations survived adjustment for inflammatory exposures, which suggests that the weak effects of SES on the neonatal proteome that we observed in a small number of EpiScores are at least partially accounted for by inflammatory pathologies in early life. Taken together, the results suggest immune dysregulation, proxied by EpiScores, may not be the primary axis through which SES becomes embedded in the development of preterm infants during neonatal intensive care.

SES has been consistently associated with inflammation in adulthood, including in relation to childhood deprivation [15,105], but less is known about the relationship between SES and inflammation in the neonatal period. A longitudinal study by Leviton *et al* [106], with five sampling timepoints during the first month of life after preterm birth, showed that maternal eligibility for Medicaid associated with levels of 14 inflammatory proteins (IL6R, TNFR1, TNFR2, IL8, ICAM1, VCAM1, TSH, EPO, bFGF, IGF1, VEGF, PIGF, Ang-1, Ang-2). However, only three were significant at more than one timepoint during the month after preterm birth (TSH, bFGF, Ang-1), and none associated at all five measurement timepoints. These studies, taken together with our results, suggest that the impact of SES on immune regulation is relatively modest and inconsistent in the newborn period but accrues through to adulthood. Further research is required to understand how and when SES becomes embedded in child development and whether early life events such as preterm birth modify that process; EpiScores could be a powerful tool for investigating the temporal dynamics of social determinants of child health.

### 4.3 Strengths and limitations

Strengths of this study include the large sample of term and preterm neonates; to the best of our knowledge, this is the first examination of multiple DNA methylation-based estimators of circulating proteins in a neonatal sample. We derived EpiScores from minimally-invasive sampling (buccal cells from saliva) which overcomes the ethical challenge of venepuncture for research in children. The EpiScores, serving as proxies of inflammatory proteins and sampled at term-equivalent age in preterm infants, were selected for their potential to capture chronic, cumulative inflammation associated with preterm birth and neonatal intensive care exposures [23–25]. We adjusted for variables associated with DNAm, and additionally for inflammatory exposures to increase the clinical validity of our results.

The study has some limitations. The EpiScores used were developed in adult cohorts [26–28,37] and have not been validated in neonatal populations with neonatal protein levels, although we have previously established that neonatal DNAmCRP scores correlate with cumulative inflammatory exposures [32]. Further studies that evaluate serial blood protein levels with EpiScores could be informative but would need to rely on small volume samples taken during time of venepuncture for clinical reasons, given the practical and ethical challenges of serial venepuncture for research purposes in neonates. The EpiScores were also trained using blood samples [26–28,37], whereas we have projected these scores into saliva samples. However, previous studies have successfully used similar cross-tissue techniques [32,107,108], and in neonates saliva provides a non-invasive and accessible sample method. Not all inflammatory-related proteins are represented, as we were limited by available EpiScores, so we may have underestimated the full complexity of the relationship between birth GA, SES, and inflammation. Longitudinal investigations are imperative for elucidating whether the DNA methylation signatures associated with gestational age identified in this study exert a causal influence on the inflammation-associated mechanisms in preterm birth. It remains crucial to discern whether these signatures represent a direct downstream consequence of GA itself or are induced by specific factors correlated with GA, yet not necessarily driven by chronic inflammation. Mendelian randomization studies, integrating genomic and epigenomic determinants, are a promising methodological approach to disentangle the directionality of these intricate relationships. The study population is comparable to other neonatal populations in high-income, majority-white settings, but these results may not generalise to settings with different socioeconomic or ethnicity profiles. We studied several measures of SES but were not able to include all that could be relevant, such as household income.

## 5.0 Conclusion

We identified 43 EpiScores enriched for inflammatory proteins that associated with low birth GA. These 43 proteins offer novel insights into the physiological response to preterm birth and warrant further study to explore their role in the relationship between preterm birth, inflammation, and longer-term outcomes. We found only three EpiScores associated with SES in the neonatal period, none of which survived adjustment for perinatal pro-inflammatory exposures, suggesting that inflammation is unlikely to be the primary axis through which SES becomes embedded in the development of preterm infants in the neonatal period.

## 6.0 Declarations

### 6.1 Ethics approval and consent to participate

Ethical approval for the study was obtained from the National Research Ethics Service, South-East Scotland Research Ethics Committee (REC 11/55/0061, 13/SS/0143, 16/SS/0154), and NHS Lothian Research and Development (2016/0255). Written informed consent was obtained from parents.

### 6.2 Consent for publication

Not applicable.

### 6.3 Availability of data and materials

DNA methylation data are available to researchers subject to the terms of the Data Access Policy: https://www.ed.ac.uk/centre-reproductive-health/tebc/about-tebc/for-researchers/data-access-collaboration.

### 6.6 Authors’ contributions

KM: Conceptualisation, methodology, formal analysis, writing (original draft), writing (review/editing), visualization. ELSC: Formal analysis, writing (review/editing). KV: Formal analysis, writing (review/editing), visualization. RH: Methodology, formal analysis, writing (review/editing). DG: Methodology, formal analysis, writing (review/editing). JB: Formal analysis, writing (review/editing). GS: Investigation, writing (review/editing). AJS: Methodology, writing (review/editing). AC: Investigation, resources, data curation, writing (review/editing). LM: Investigation, resources, writing (review/editing). HCW: Methodology, writing (review/editing). HR: Conceptualisation, writing (original draft), writing (review/editing), supervision. REM: Conceptualisation, methodology, writing (original draft), writing (review/editing), supervision. SRC: Conceptualisation, methodology, writing (original draft), writing (review/editing), supervision. JPB: Conceptualisation, methodology, writing (original draft), writing (review/editing), supervision, project administration, funding acquisition.

## 6.7 Acknowledgements

The authors are grateful to the families who consented to take part in the study.

