## Additional File 1 for "Epigenetic scores indicate differences in the proteome of preterm infants"

**Supplementary information**

Katie Mckinnon, Eleanor L.S. Conole, Kadi Vaher, Robert F. Hillary, Danni A. Gadd, Justyna Binkowska, Gemma Sullivan, Anna J. Stevenson, Jill Hall, Amy Corrigan, Lee Murphy, Heather C. Whalley, Hilary Richardson, Riccardo E. Marioni, Simon R. Cox, James P. Boardman

**Contents**

Supplementary eMethods

Supplementary Table 1. Spreadsheet of EpiScores with CpG sites and coefficients

Supplementary Table 2. Frequency of perinatal pro-inflammatory co-exposures

Supplementary Table 3. Standardised component loading of the two principal components

Supplementary Figure 1. Relationship between EpiScores and birth gestational age in regression models with the Scottish Index of Multiple Deprivation as socioeconomic status measure

Supplementary Figure 2. Relationship between EpiScores and an interaction between birth gestational age and socioeconomic status in regression models with the Scottish Index of Multiple Deprivation as socioeconomic status measure

Supplementary Figure 3. Relationship between EpiScores and the Scottish Index of Multiple Deprivation as socioeconomic status measure

Supplementary Figure 4. Relationship between EpiScores and birth gestational age in regression models with maternal education as socioeconomic status measure

Supplementary Figure 5. Relationship between EpiScores and an interaction between birth gestational age and socioeconomic status in regression models with maternal education as socioeconomic status measure

Supplementary Figure 6. Relationship between EpiScores and maternal education as socioeconomic status measure

Supplementary Figure 7. Relationship between EpiScores and birth gestational age in regression models with maternal occupation as socioeconomic status measure

Supplementary Figure 8. Relationship between EpiScores and interaction between birth gestational age and socioeconomic status in regression models with maternal occupation as socioeconomic status measure

Supplementary Figure 9. Relationship between EpiScores and maternal occupation as socioeconomic status measure

Supplementary Table 4. Functional roles of proteins of EpiScores associated with birth gestational age

Supplementary Figure 10. Interaction between afamin EpiScore, birth gestational age, and maternal education

Supplementary Figure 11. Interaction between HGFI EpiScore, birth gestational age, and maternal occupation

Supplementary Table 5. Relationship between EpiScores and socioeconomic status measure in unadjusted and adjusted models

**eMethods**

Predictor variables:

- Neighbourhood-level SES. Scottish Index of Multiple Deprivation (SIMD, 2016) was derived from the family’s postcode at time of birth[1] (n=331).
  - Continuous measure, from 1 (most deprived) to 6976 (least deprived). SIMD was chosen because it incorporates multiple dimensions (population characteristics, income, employment, health, education, access to services, crime, and housing)[1], neighbourhood deprivation is consistently associated with child development[2], and SIMD is a tractable tool for policy makers[1].
- Family-level SES. Collected from questionnaires at recruitment.
  - Maternal education: highest educational qualification, as ordered factor measure (n=323).
    - None: no qualifications obtained.
    - Basic high school qualification: National 5s, Standard Grades, GCSEs (General Certificates of Secondary Education) or equivalent. This is divided into 1-4 and >5 qualifications.
    - Advanced high school qualification: Highers, A levels (Advanced levels) or equivalent.
    - College qualification: e.g. National Certificate, Higher National Diploma, Higher National Certificate, vocational qualifications.
    - University undergraduate degree.
    - University postgraduate degree.
  - Maternal occupation: current/most recent occupation, as a factor measure (n=328).
    - Unemployed.
    - Homemaker.
    - Still in full time education.
    - Unskilled: cleaners, porters, messengers.
    - Partly skilled: e.g. bus conductor, postman.
    - Manual skilled: e.g. toolmaker, foreman, ambulance man.
    - Non-manual skilled: e.g. typist, police officer, fireman.
    - Professional: e.g. doctors, lawyers, teachers, managers.

Pro-inflammatory covariates:

- Histological chorioamnionitis (HCA), from placental histology.
- Sepsis, from medical records. Positive blood culture and/or physician decision to treat with five days of antibiotics; early-onset is <72 hours after birth and late-onset is after 72 hours.
- Bronchopulmonary dysplasia (BPD), from medical records. Requirement for respiratory support and/or supplemental oxygen after 36 weeks' gestation.
- Necrotising enterocolitis (NEC), from medical records. Medical (7 days nil by mouth) or surgical management.

EpiScores from Gadd *et al*[3] not included here:

- EpiScores removed due to unavailable cytosine-phosphate-guanine dinucleotide (CpGs): BCAM, Lysozyme C, MMP2.
- EpiScores removed due to duplication: CXCL10, CXCL11, MMP1, Granzyme A, NTRK3.

**Supplementary Table 1.** **Spreadsheet of EpiScores with CpG sites and coefficients**

Excel spreadsheet: Supplementary_Table_1.xlsx

104 included EpiScores with CpG sites and coefficients required to calculate scores for each individual.

**Supplementary Table 2. Frequency of perinatal pro-inflammatory co-exposures**

| **Exposure** | **Preterm (total n=217)** |
| --- | --- |
| Histological chorioamnionitis | 61/158 (38.6%) |
| Sepsis | 56/217 (25.8%) |
| Bronchopulmonary dysplasia | 66/215 (30.7%) |
| Necrotising enterocolitis | 11/217 (5.5%) |

**Supplementary Table 3. Standardised component loading of the two principal components**

| **EpiScore** | **PC 1** | **PC 2** |
| --- | --- | --- |
| ADAMTS | 0.042 | 0.081 |
| Adiponectin | 0.055 | -0.095 |
| Afamin | -0.169 | -0.002 |
| Alpha-L-iduronidase | 0.029 | 0.035 |
| Aminoacylase-1 | -0.162 | -0.004 |
| Beta-2-microglobulin | 0.068 | -0.123 |
| BMP1 | -0.156 | 0.104 |
| CCL11 | 0.005 | -0.164 |
| CCL17 | -0.046 | 0.029 |
| CCL18 | 0.035 | 0.074 |
| CCL21 | -0.034 | -0.064 |
| CCL22 | -0.048 | -0.144 |
| CCL25 | -0.112 | 0.037 |
| CD163 | -0.101 | -0.111 |
| CD209 | 0.191 | -0.039 |
| CD48 | -0.099 | -0.081 |
| CD5L | 0.094 | -0.162 |
| CD6 | -0.085 | -0.190 |
| CHIT1 | 0.019 | -0.193 |
| Coagulation factor VII | -0.003 | -0.002 |
| Complement C4 | 0.044 | 0.044 |
| Complement C5a | -0.163 | -0.096 |
| Complement c9 | -0.071 | 0.045 |
| Contactin-4 | 0.088 | 0.114 |
| CRP | -0.138 | 0.013 |
| CRTAM | 0.107 | 0.020 |
| CXCL10 | 0.058 | -0.024 |
| CXCL11 | -0.033 | -0.108 |
| CXCL9 | 0.029 | 0.086 |
| E-selectin | -0.068 | 0.062 |
| Ectodysplasin-A | 0.039 | -0.093 |
| ENPP7 | -0.096 | -0.112 |
| ESM1 | 0.110 | -0.064 |
| Ezrin | 0.108 | -0.188 |
| FAP | 0.092 | 0.008 |
| FCER2 | -0.097 | -0.125 |
| FCGR3A | -0.080 | -0.063 |
| FcRL2 | -0.127 | -0.194 |
| FGF21 | -0.050 | -0.113 |
| Galectin-4 | 0.142 | -0.092 |
| GCSF | 0.106 | -0.005 |
| GDF15 | -0.040 | 0.106 |
| GDF8 | 0.168 | -0.044 |
| GHR | -0.009 | -0.087 |
| GPIba | 0.181 | -0.040 |
| Granulysin | -0.033 | -0.138 |
| Granzyme A | 0.065 | -0.191 |
| HCII | 0.021 | 0.055 |
| HGF | -0.055 | 0.020 |
| HGFA | -0.160 | 0.097 |
| HGFI | -0.002 | -0.006 |
| ICAM5 | 0.132 | -0.087 |
| IGFBP1 | 0.081 | -0.010 |
| IGFBP4 | -0.115 | -0.130 |
| IL19 | -0.064 | -0.073 |
| IL6 | 0.009 | -0.167 |
| Insulin receptor | -0.102 | -0.143 |
| L-selectin | 0.172 | 0.136 |
| Lactotransferrin | 0.022 | 0.031 |
| LGALS3BP | -0.032 | -0.110 |
| LY9 | -0.030 | -0.039 |
| Lymphotoxin | 0.131 | -0.068 |
| MIA | -0.036 | -0.125 |
| MMP1 | -0.176 | 0.007 |
| MMP12 | -0.026 | -0.141 |
| MMP9 | -0.056 | 0.086 |
| MRC2 | -0.107 | -0.086 |
| Myeloperoxidase | -0.112 | 0.092 |
| NCAM1 | 0.151 | -0.108 |
| NCDase | 0.022 | 0.004 |
| Neprilysin | 0.009 | -0.106 |
| NMNAT1 | 0.171 | -0.037 |
| NOTCH1 | 0.128 | 0.030 |
| NTproBNP | -0.057 | 0.136 |
| NTRK3 | 0.022 | 0.152 |
| Oncostatin-M | 0.166 | 0.046 |
| Osteomodulin | -0.027 | 0.037 |
| Pappalysin-1 | 0.114 | 0.060 |
| PIGR | -0.019 | -0.033 |
| RARRES2 | 0.147 | -0.043 |
| Resistin | 0.155 | 0.117 |
| S100A12 | 0.129 | 0.109 |
| S100A9 | 0.109 | 0.179 |
| SCGF-alpha | -0.050 | 0.180 |
| SCGF-beta | -0.088 | 0.159 |
| Semaphorin-3E | 0.081 | -0.076 |
| SERPINA3 | 0.032 | <0.001 |
| SHBG | 0.123 | -0.032 |
| Sialoadhesin | -0.111 | -0.075 |
| SKR3 | -0.013 | 0.002 |
| SLITRK5 | 0.169 | -0.047 |
| SMPD1 | -0.068 | -0.010 |
| Stanniocalcin-1 | -0.092 | 0.010 |
| Testican-2 | 0.013 | -0.134 |
| TGF alpha | 0.070 | 0.132 |
| Thrombopoietin receptor | 0.164 | -0.075 |
| Thrombospondin-2 | -0.007 | -0.102 |
| TNFRSF17 | -0.156 | 0.108 |
| TNFRSF1B | 0.079 | -0.203 |
| Trypsin-2 | -0.020 | 0.021 |
| Tryptase beta-2 | 0.077 | -0.007 |
| VCAM1 | -0.041 | -0.090 |
| VEGFA | -0.118 | -0.040 |
| WFIKKN2 | 0.097 | -0.026 |

Component loadings for the 104 EpiScores for the two principal components.

PC – principal component

**Supplementary Figure 1. Relationship between EpiScores and birth gestational age in regression models with the Scottish Index of Multiple Deprivation as socioeconomic status measure**

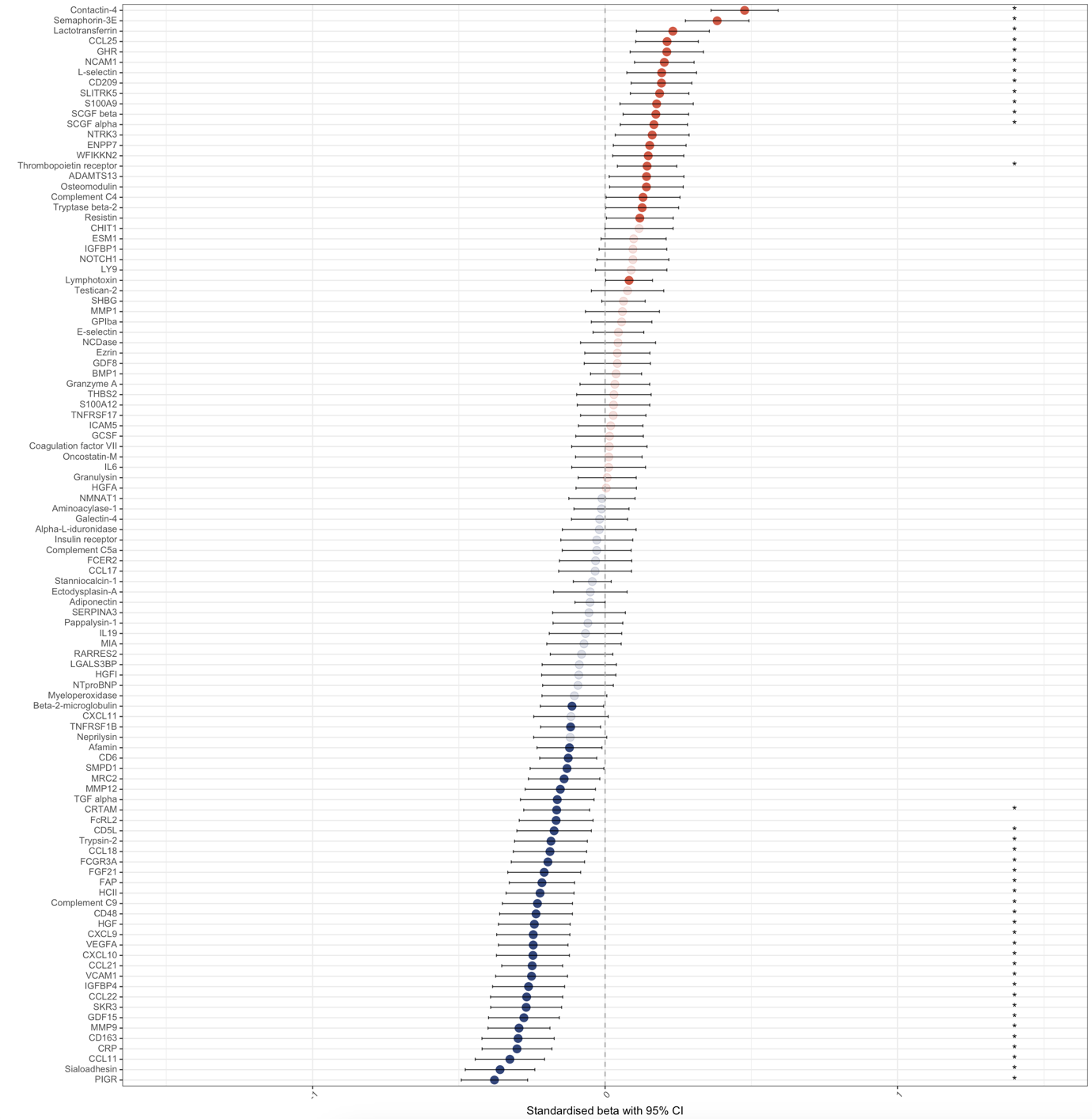

EpiScores associated with birth gestational age in regression models with the Scottish Index of Multiple Deprivation, gestational age at sample, birthweight z-score, sex, and methylation processing batch. n=331. Standardised beta with 95% confidence intervals. Red are positive associations, blue are negative associations. Those with strong colour are individually significant with *p*<0.05 (57/104 EpiScores), and those with an asterisk are significant at the adjusted *p*<8.3x10^-3^ (39/104 EpiScores).

CI – confidence interval

**Supplementary Figure 2. Relationship between EpiScores and an interaction between birth gestational age and socioeconomic status in regression models with the Scottish Index of Multiple Deprivation as socioeconomic status measure**

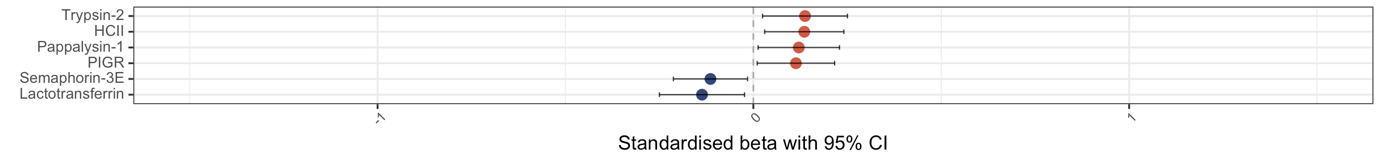

EpiScores associated with an interaction between birth gestational age and the Scottish Index of Multiple Deprivation in regression models with the Scottish Index of Multiple Deprivation, gestational age at birth, gestational age at sample, birthweight z-score, sex, and methylation processing batch. n=331. Standardised beta with 95% confidence intervals. Red are positive associations, blue are negative associations. Those with strong colour are individually significant with *p*<0.05 (6/104 EpiScores), and those with an asterisk are significant at the adjusted *p*<8.3x10^-3^ (0/104 EpiScores).

CI – confidence interval

**Supplementary Figure 3. Relationship between EpiScores and the Scottish Index of Multiple Deprivation as socioeconomic status measure**

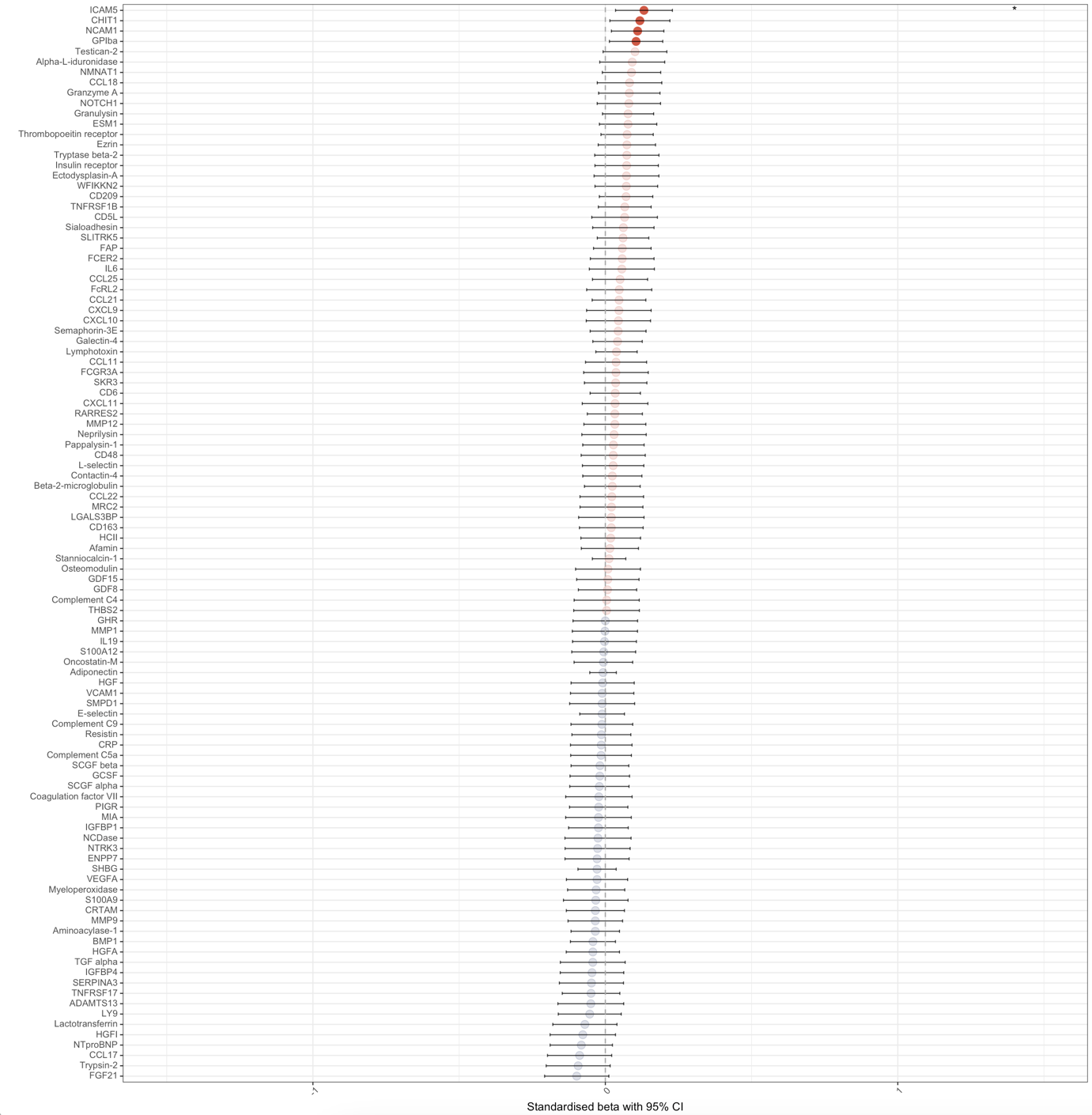

EpiScores associated with the Scottish Index of Multiple Deprivation in regression models with gestational age at birth, gestational age at sample, birthweight z-score, sex, and methylation processing batch. n=331. Standardised beta with 95% confidence intervals. Red are positive associations, blue are negative associations. Those with strong colour are individually significant with *p*<0.05 (4/104 EpiScores), and those with an asterisk are significant at the adjusted *p*<8.3x10^-3^ (1/104 EpiScores).

CI – confidence interval

**Supplementary Figure 4. Relationship between EpiScores and birth gestational age in regression models with maternal education as socioeconomic status measure**

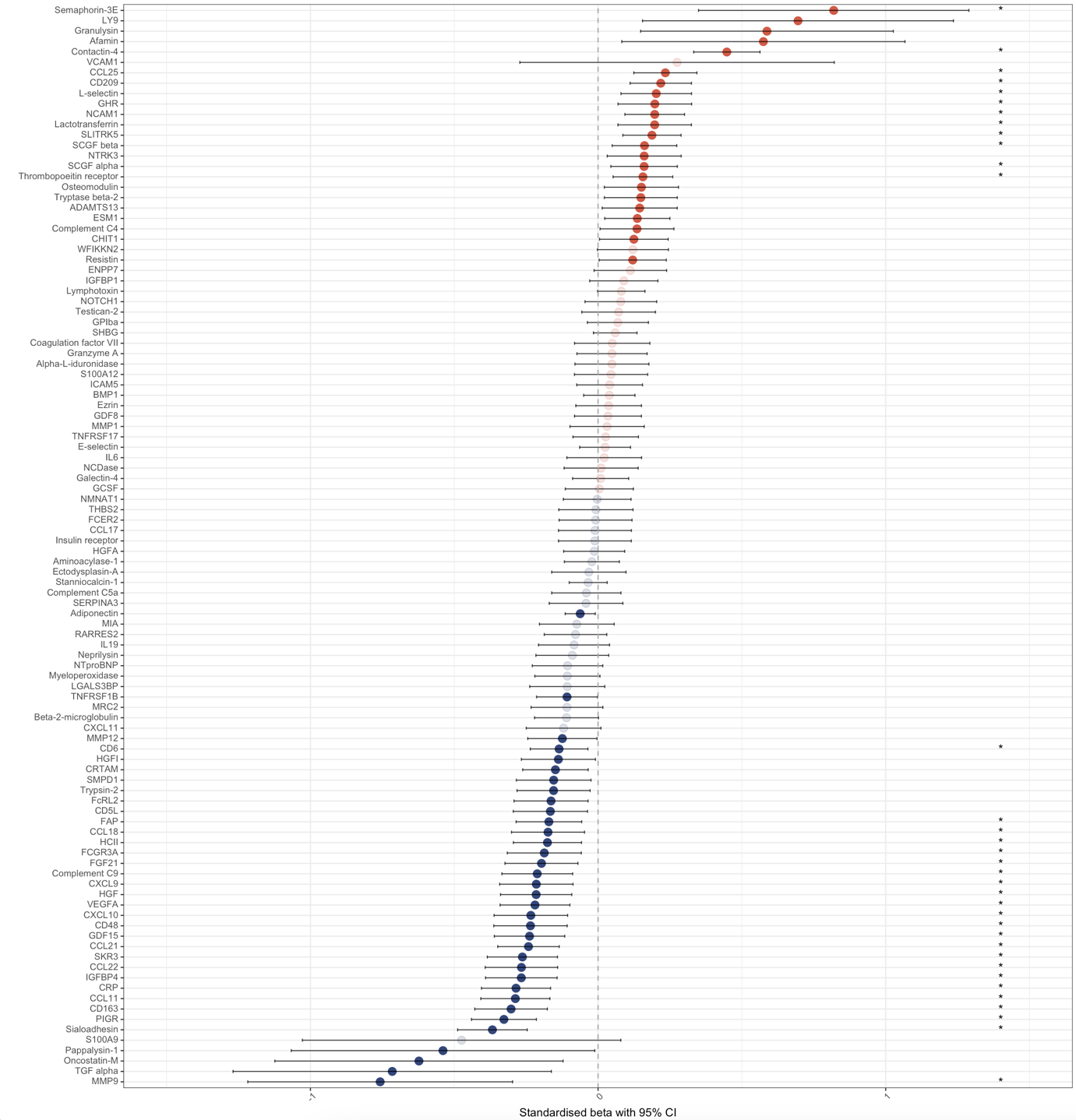

EpiScores associated with birth gestational age in regression models with maternal education, gestational age at sample, birthweight z-score, sex, and methylation processing batch. n=323. Standardised beta with 95% confidence intervals. Red are positive associations, blue are negative associations. Those with strong colour are individually significant with *p*<0.05 (58/104 EpiScores), and those with an asterisk are significant at the adjusted *p*<8.3x10^-3^ (35/104 EpiScores).

CI – confidence interval

**Supplementary Figure 5. Relationship between EpiScores and an interaction between birth gestational age and socioeconomic status in regression models with maternal education as socioeconomic status measure**

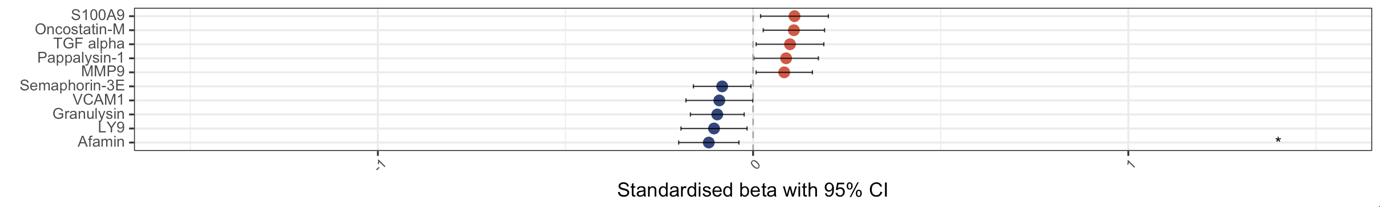

EpiScores associated with an interaction between birth gestational age and maternal education in regression models with maternal education, gestational age at birth, gestational age at sample, birthweight z-score, sex, and methylation processing batch. n=323. Standardised beta with 95% confidence intervals. Red are positive associations, blue are negative associations. Those with strong colour are individually significant with *p*<0.05 (10/104 EpiScores), and those with an asterisk are significant at the adjusted *p*<8.3x10^-3^ (1/104 EpiScores).

CI – confidence interval

**Supplementary Figure 6. Relationship between EpiScores and maternal education as socioeconomic status measure**

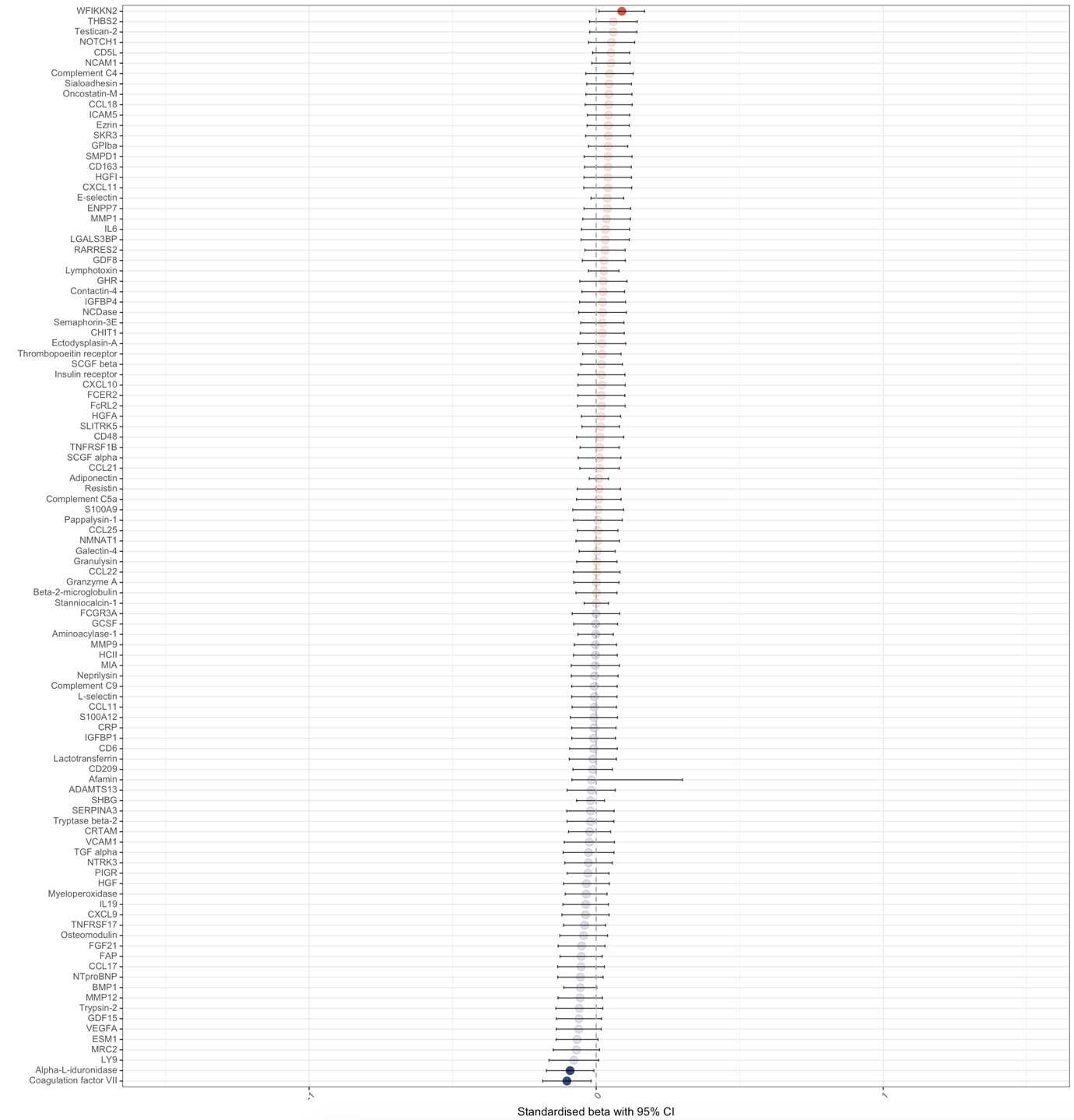

EpiScores associated with maternal education in regression models with the gestational age at birth, gestational age at sample, birthweight z-score, sex, and methylation processing batch. n=323. Standardised beta with 95% confidence intervals. Red are positive associations, blue are negative associations. Those with strong colour are individually significant with *p*<0.05 (3/104 EpiScores), and those with an asterisk are significant at the adjusted *p*<8.3x10^-3^ (0/104 EpiScores).

CI – confidence interval

**Supplementary Figure 7. Relationship between EpiScores and birth gestational age in regression models with maternal occupation as socioeconomic status measure**

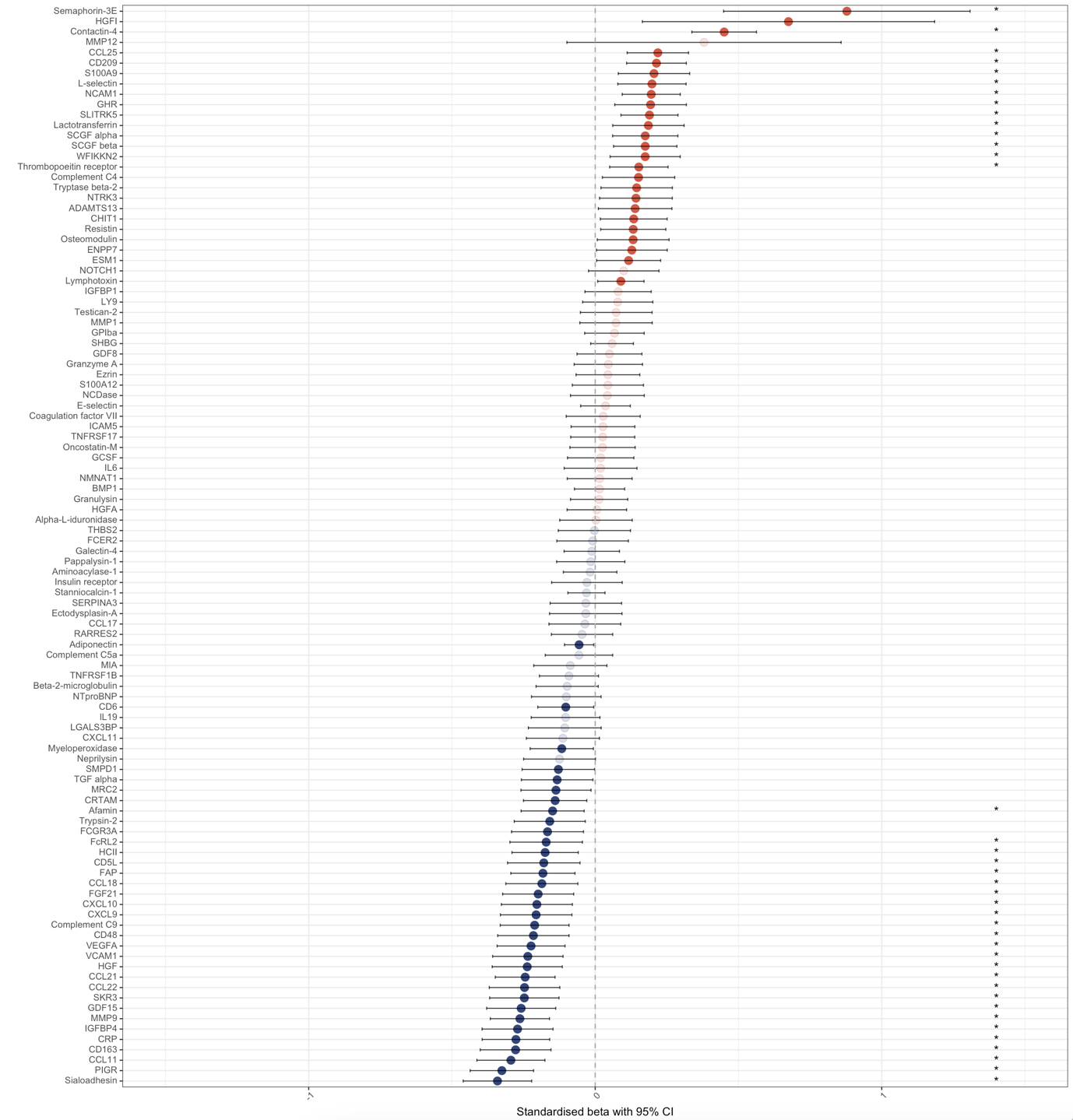

EpiScores associated with birth gestational age in regression models with maternal occupation, gestational age at sample, birthweight z-score, sex, and methylation processing batch. n=328. Standardised beta with 95% confidence intervals. Red are positive associations, blue are negative associations. Those with strong colour are individually significant with *p*<0.05 (59/104 EpiScores), and those with an asterisk are significant at the adjusted *p*<8.3x10^-3^ (39/104 EpiScores).

CI – confidence interval

**Supplementary Figure 8. Relationship between EpiScoresan interaction between birth gestational age and socioeconomic status in regression models with maternal occupation as socioeconomic status measure**

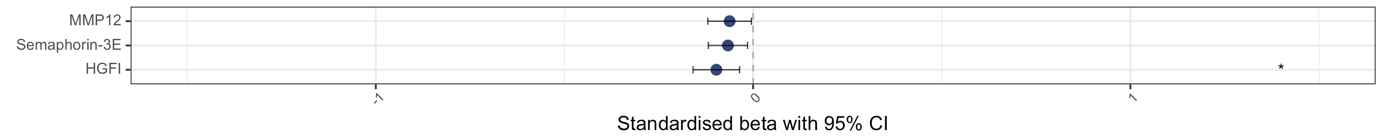

EpiScores associated with an interaction between birth gestational age and maternal occupation in regression models with maternal occupation, gestational age at birth, gestational age at sample, birthweight z-score, sex, and methylation processing batch. n=328. Standardised beta with 95% confidence intervals. Red are positive associations, blue are negative associations. Those with strong colour are individually significant with *p*<0.05 (3/104 EpiScores), and those with an asterisk are significant at the adjusted *p*<8.3x10^-3^ (1/104 EpiScores).

CI – confidence interval

**Supplementary Figure 9. Relationship between EpiScores and maternal occupation as socioeconomic status measure**

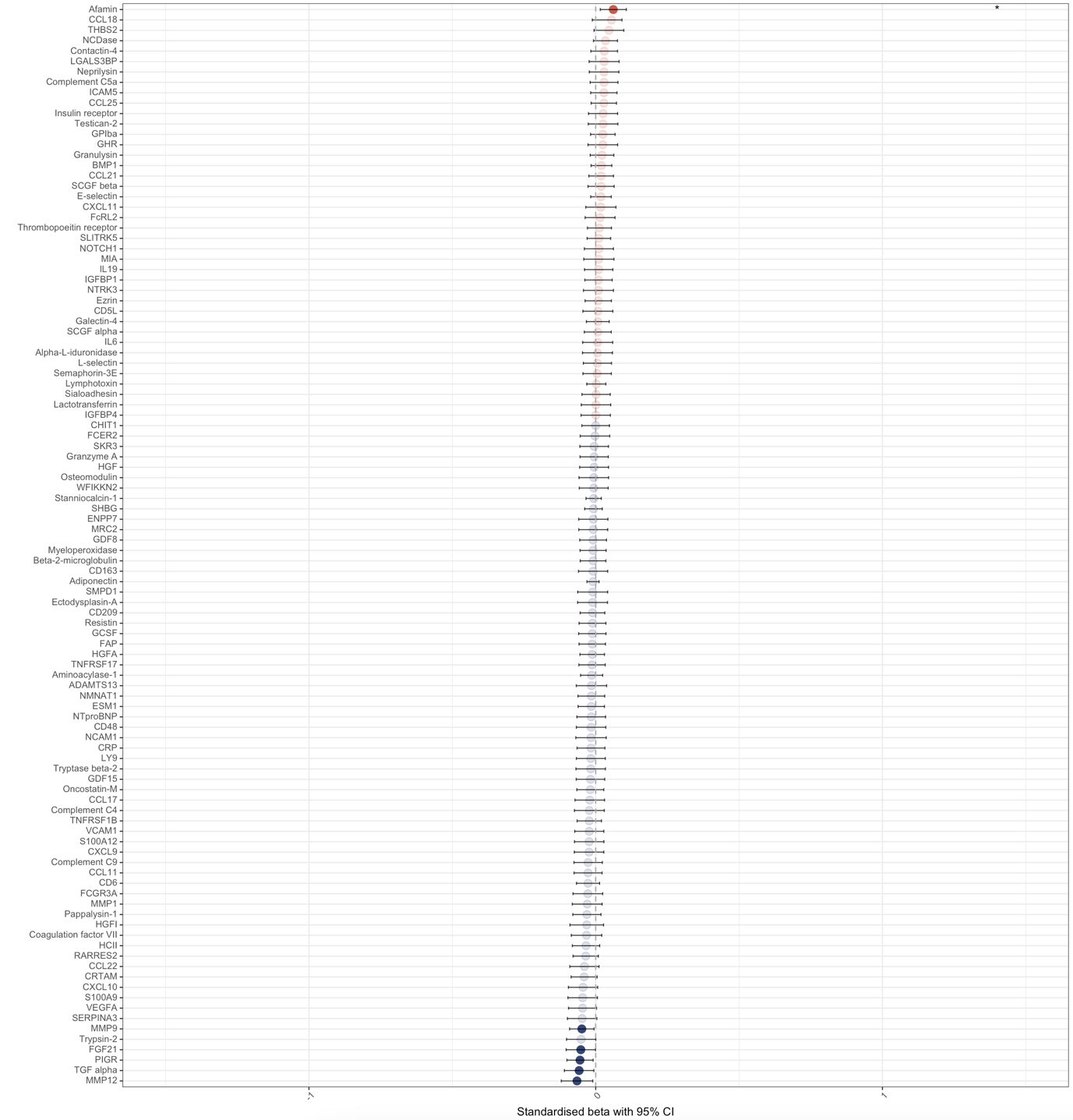

EpiScores associated with maternal occupation in regression models with gestational age at birth, gestational age at sample, birthweight z-score, sex, and methylation processing batch. n=328. Standardised beta with 95% confidence intervals. Red are positive associations, blue are negative associations. Those with strong colour are individually significant with *p*<0.05 (6/104 EpiScores), and those with an asterisk are significant at the adjusted *p*<8.3x10^-3^ (1/104 EpiScores).

CI – confidence interval

**Supplementary Table 4. Functional roles of proteins of EpiScores associated with birth gestational age**

| **EpiScore protein** | **Function and associations** | **Association with birth gestational age** |
| --- | --- | --- |
| Afamin (AFM) | Afamin is a vitamin E binding protein, involved in the transport of vitamin E in body fluids, including across the blood-brain-barrier[4,5], and anti-inflammatory. Afamin EpiScore is associated with diabetes[3], and in pregnant women it afamin is increased with pre-eclampsia and gestational diabetes[6]. In preterm infants, increased afamin is associated with BPD[7]. | Negative |
| CCL11 (C-C motif chemokine 11, eotaxin) | A chemokine involved in the stimulation of eosinophils. CCL11 is involved in allergic responses, but also in psychiatric disorders and Alzheimer’s disease[8–10]. In cord blood, CCL11 concentration is increased with GA[11], and in the presence of vasculopathy and inflammation[12]. CCL11 concentration is increased in blood in infants with BPD[13], patent ductus arteriosus (PDA)[14], and ROP[15], and vitreous fluid in ROP[16]. CCL11 EpiScore is associated with depression and cardiovascular disease in adults[3,17]. | Negative |
| CCL18 (C-C motif chemokine 18) | CCL18 is an anti-inflammatory chemokine, involved in T-cell recruitment and the regulation of other chemokines[18,19]. CCL18 increases after birth in preterm infants[20], and lower CCL18 is associated with intraventricular haemorrhage and cerebral palsy[21,22]. | Negative |
| CCL21 (C-C motif chemokine 21) | CCL21 stimulates immune cell migration and angiogenesis, with a key role in joint inflammation in rheumatoid arthritis[23]. CCL21/CCR7 signalling is involved in the regulation of neuroinflammation[24]. CCL21 is significantly increased in adults and preterm infants with sepsis.[25,26] | Negative |
| CCL22 (C-C motif chemokine 22, MDC, macrophage-derived chemokine) | CCL22 is a macrophage-derived chemokine, expressed in the thymus and chemotactic for monocytes, T-cells, and NK-cells[27]. CCL22 plays a role in neuroinflammation; anti-CCL22 reduces inflammation in an animal model of multiple sclerosis (MS)[28], and CCL22 is raised in adults with first-episode psychosis and correlates with symptoms[29]. CCL22 is involved in allergic conditions[27], and mediates inflammation in haemorrhage-induced lung injury[30]. In an animal model, clarithromycin to prevent preterm birth reduces CCL22 levels.[31] | Negative |
| CCL25 (C-C motif chemokine 25, TECK, thymus-expressed chemokine) | CCL25 is involved in immune function in the thymus, intestine, heart, joints and trachea, and involved in a range of adult inflammatory conditions such as inflammatory bowel disease (IBD), asthma and ischaemic heart disease (IHD)[32]. CCL25 is present in breast milk, particularly colostrum[33]. CCL25 is different between mothers with threatened preterm labour, and those with term infants, although not with those with preterm infants[34]. CCL25 is increased in adults with sepsis, and anti-CCL25 reduces inflammation and lung injury in a mouse model of sepsis[26]. | Positive |
| CD5L (CD5 antigen-like, AIM) | CD5L has multiple anti-inflammatory roles, particularly in the regulation of leukocyte function such as apoptosis inhibition[35]. It is involved in responses to various bacterial infections[35], has been suggested as a treatment for sepsis[36], and is altered by prophylactic antibiotic treatment in an animal model of preterm birth[37]. It has also reduced inflammation in an animal model of ischaemic stroke[38]. | Negative |
| CD6 (T-cell differentiation antigen) | CD6 is a lymphocyte receptor that binds to gram-positive bacteria through lipoteichoic acid, and gram-negative bacteria through lipopolysaccharides (LPS)[39]. CD6 is involved in the early immune response to bacterial sepsis, through immune cell proliferation and function[40], although CD6 is downregulated in sepsis[41]. CD6 also plays a role in auto-immune conditions, such as psoriasis, rheumatoid arthritis, and MS[42]. | Negative |
| CD48 (CD48 antigen) | CD48 is expressed on T-cells and antigen-presenting cells with immune functions[43], and expressed on haematopoeitic stem cells (HSCs) for regulation of stem cells and tumour suppression[44]. It may have roles in allergic airway inflammation, IBD, and bacterial infections[45]. Hypoxia reduced the number of CD48+ HSCs in a mouse model of lung injury[46]. CD48 expression is increased in idiopathic inflammatory myopathies[47]. The presence of CD48 in amniotic fluid is predictive of preterm birth[48], and CD48 gene expression is increased in umbilical cord tissue from preterm infants[49]. | Negative |
| CD163 (Scavenger receptor cysteine-rich type 1 protein M130) | An acute-phase regulated receptor on macrophages, acting as a scavenger-receptor for haemoglobin-haptoglobin complexes, with specific association with a range of inflammatory diseases such as bacterial sepsis, rheumatoid arthritis and IBD[50,51]. CD163 positive cells are higher in placentas from preterm than term infants[52], and in the blood and small intestine mucosa in infants with NEC[53]. In neonates and children with sepsis, those with systemic inflammatory response syndrome have increased expression of CD163[54], and in an animal model, CD163 is involved in PDA closure[55]. CD163 EpiScore was associated with diabetes in adults[3]. | Negative |
| CD209 (DC-SIGN, dendritic cell-specific ICAM-grabbing nonintegrin) | CD209 is expressed on monocyte-derived macrophages, interacting with intercellular adhesion molecules and involved in T-cell mediated immunity[56]. CD209+ cells are present in the decidua during pregnancy[57], and expression in reduced in mothers with spontaneous preterm labour[58]. CD209+ dendritic cells are also increased in inflamed joints in rheumatoid arthritis and psoriatic arthritis, with increased cytokine production[59]. | Positive |
| Complement C9 (CC9) | Complement C9 forms part of the terminal complement complex (TCC) with complement components C5b-9, the membrane attack complex (MAC). TCC is increased in preterm infants in the presence of early onset sepsis[60], and complement C9 is increased in cord blood in the presence of HCA[61]. Deficiencies in C5b-9 complement components increase the risk of certain specific infections such as Neisseria, Haemophilus influenza type B and meningococcus[62]. Complement C9 administration improved survival with Ecoli sepsis in a rat model[63]. However, complement C9 is low in newborn infants compared to adults, increasing after birth from fetal levels[64,65]. Complement C9 expression may be increased in pregnancies leading to spontaneous preterm birth[66], but also rises in labour regardless of prematurity[67]. In term infants with hypoxic ischaemic encephalopathy, cerebrospinal fluid (CSF) complement C9 is reduced and TCC increased, and complement C9 deposition is found across the brain tissues on histopathology[68], and higher TCC levels in CSF correlates with abnormal developmental outcomes[69]. | Negative |
| Contactin-4 (CNTN4, brain-derived immunoglobulin superfamily protein 2, BIG2) | Contactin-4 is an immunoglobulin involved in axon guidance and neurite promotion[70,71]. It is expressed across the central nervous system (CNS), but also in the testis, thyroid, small intestine, and uterus[70,71]. Contactin-4 can also modulate amyloid precursor protein[71]. Contactin-4 increases between birth and term corrected GA in preterm infants[20]. The contactin-4 gene has been suggested as a susceptibility locus for autism[72,73], as well as neuropsychiatric conditions such as anorexia nervosa, Alzheimer’s disease, and schizophrenia[74], and a knock-out mouse model has shown changes in hippocampal synaptic development and changes to memory and stress responses[74]. | Positive |
| CRP (C-reactive protein) | An acute phase inflammatory protein, with actions through activation of the complement pathway, phagocytic cells, and affecting antigen presentation[75]. CRP is increased in preterm infants compared to term infants in cord blood[76] and in blood in the days after birth[77]. Higher concentrations of CRP in the first weeks of life are associated with ventriculomegaly[78], and increased likelihood of executive dysfunction and attention-deficit hyperactivity disorder (ADHD) symptoms at 10 years of age[79,80], although not cerebral palsy[81]. CRP EpiScore is associated with brain structures in adults, in cognitive aging and depression[82,83], as well as with mental health in childhood[84] and psychotic episodes in adulthood[85], and IHD and chronic obstructive pulmonary disease (COPD) in adults[3]. CRP EpiScore is associated with gestational age, perinatal inflammatory processes and neonatal brain development on MRI at term-corrected gestation[86]. | Negative |
| CRTAM (Cytotoxic and regulatory T-cell molecule, Class-I MHC-Restricted T-Cell-Associated Molecule) | CRTAM is an immunoglobulin-superfamily transmembrane protein, expressed on CD4+ and CD8+ T cells, NK cells and NKT cells, playing a role in cellular adhesion, polarity, proliferation and cytokine release[87]. CRTAM is important in intestinal immune responses to parasitic infections particularly[88,89], and its role in other intestinal infections may be related to CRTAM shaping the gut microbiome[90]. | Negative |
| CXCL9 (C-X-C motif chemokine 9, monokine induced by interferon gamma, MIG) | CXCL9 is a chemokine involved in immune cell movement and activation, predominantly regulated by interferon gamma. CXCL9 is involved in the acute and chronic inflammatory response to parasitic infection[91], and raised in fungal infections[25]. In pregnant women with malaria, high CXCL9 levels are associated with pregnancy loss and preterm birth[92]. CXCL9 is low in in women with premature rupture of membranes (PROM)[93], and women with rhesus D alloimmunisation[94], but raised in the mother and placenta in chorioamnionitis[95]. CXCL9 is involved in neuroinflammation, such as MS and the inflammatory response to CNS infections[96]. CXCL9 is also involved in adult inflammatory lung processes such as adenocarcinoma and interstitial lung diseases[97,98]. | Negative |
| CXCL10 (C-X-C motif chemokine 10, interferon gamma-inducible protein 10, IP10) | CXCL10 is chemotactic for monocytes and T-lymphocytes, with a particular role in neuroinflammation, for example MS, Alzheimer’s disease and CNS infections[96,99]. CXCL10 is increased in amniotic fluid in the presence of chronic chorioamnionitis[100], and in mothers with preterm labour[101]. CXCL10 is also involved in the acute and chronic inflammatory response to parasitic infection[91], and interstitial lung disease[98]. In a neonatal animal model, hyperglycaemia stimulates CXCL10[102], and in preterm endothelial cells there is reduced CXCL10 production following pro-inflammatory stimulation compared to term endothelial cells[103]. CXCL10 is low in in women with PROM[93], and women with rhesus D alloimmunisation[94]. In cord blood, increased CXCL10 is associated with an increased likelihood of autism[104]. CXCL10 EpiScore is associated with rheumatoid arthritis in adults[3]. | Negative |
| FAP (Fibroblast activation protein alpha, seprase) | FAP is a type-2 transmembrane serine protein, expressed on fibroblasts, melanocytes, macrophages, and in various malignant cell types[105]. FAP is involved in wound healing, liver and lung fibrosis, osteoarthritis, rheumatoid arthritis, IHD, and a range of malignancies[105]. It inactivates FGF21, so may also be involved in metabolic conditions[105]. FAP is increased in placentas with pre-eclampsia[106], and in the cord blood of preterm neonates after gestational diabetes[107]. FAP increases then decreases after birth in preterm infants[20]. | Negative |
| FCGR3B (Low affinity immunoglobulin gamma Fc region receptor III-B, CD16b) | FCGR3B is a binding site for polymeric immunoglobulin and immune complexes on neutrophils, eosinophils and basophils[108]. FCGR3B is significantly increased in preterm infants with sepsis[25], and in the placenta in chorioamnionitis[95]. The presence of FCGR3B in amniotic fluid is predictive of preterm birth[48]. FCGR3B is involved in systemic sclerosis and systemic lupus erythematosus (SLE)[109], rheumatoid arthritis[110], ulcerative colitis[111], glomerulonephritis[112], and susceptibility to malaria[113]. | Negative |
| FcRL2 (Fc receptor-like protein 2) | FcRL2 is a receptor for the constant region of immunoglobulins, inhibiting B-cell receptor activation and affecting the circulating levels of immune complexes[114,115]. FcRL2 expression is reduced in MS patients with increased neurodegeneration[116]. FcRL2 expression correlates with serum inflammatory markers and disease activity in rheumatoid arthritis[117], and is a prognostic marker in chronic lymphocytic leukaemia[118]. In children with obesity, FcRL2 expression positively correlates with muscle strength[119]. | Negative |
| FGF21 (Fibroblast growth factor 21) | FGF21 is a hormone involved in various metabolic pathways, particularly within skeletal muscle, including acting as a pro-inflammatory cytokine, correlating with other inflammatory proteins such as GDF15 and IL6[120]. FGF21 expression is increased following preterm birth[20], and raised levels are associated with postnatal growth failure[121]. FGF21 is also associated with fetal death or preterm birth in pregnancies with fetal growth restriction[122]. | Negative |
| GDF15 (Growth and differentiation factor 15) | GDF15 is also known as macrophage inhibitory cytokine 1 (MIC-1), part of the transforming growth factor beta (TGF$\beta$)[123,124]. GDF15 is secreted by the placenta, with higher levels with lower gestational age at birth[125], and levels drop with postnatal age in extremely preterm infants, particularly over the first week of life[20,125,126]. GDF15 also correlates with NTproBNP and lactate in this first week of life[126]. It has been associated with adult chronic inflammatory processes in animal models, including atherosclerosis and rheumatoid arthritis.[123] Other predominantly adult disease links include sepsis[127], pre-eclampsia[128], mitochondrial disorders (this study also included children)[129], and Alzheimer’s[130]. In preterm infants, GDF15 has particularly been linked to BPD[131,132]. Higher GDF15 levels are associated with a longer need for mechanical ventilation, prolonged respiratory support need, and length of hospital stay[125]. It has even been suggested that sex differences in GDF15 activation by hyperoxia could explain sex differences in BPD rates, although only in animal models[133]. Other relevant associations are to PDAs in preterm infants[134], and pulmonary hypertension in children[135]. It decreases between birth and term corrected GA in preterm infants[20]. | Negative |
| GHR (Growth hormone receptor) | The primary role of GHR is related to growth and metabolism[136]. GHR is correlated with body mass index (BMI)[137]. Preterm infants with the d3-variant of GHR have increased postnatal catch-up growth[138]. GHR is also present across all body tissues, including throughout the brain, and has a role in neuronal and glial proliferation and differentiation[139]. GH is also protective in animal models of hypoxic ischaemic encephalopathy[139]. GH also mediates pain in a neonatal animal model[140]. GHR knockout mice have increased longevity, and reduced inflammatory cytokines[141], with GH having other roles in inflammatory modulation such as macrophage activation[142]. | Positive |
| HCII (Heparin cofactor 2, HEP2, SERPIND1) | HCII is a serine proteinase inhibitor, inactivating thrombin, particularly in the presence of dermatan sulphate. Levels are low in term infants compared to adults, and even lower in preterm infants[143]. HCII is involved in host defence against gram-negative bacteria and in wound response[144,145]. High HCII levels are protective against in-stent restenosis and atherosclerosis[146]. HCII also plays a role in glucose homeostasis, with HCII levels negatively correlated with HbA1c and insulin resistance[147], and HCII is increased in the cord blood of preterm neonates after gestational diabetes[107]. HCII expression is increased in pregnancies leading to spontaneous preterm birth[66]. | Negative |
| HGF (Hepatocyte growth factor) | HGF mediates inflammation through actions on various cells, and their antigens and cytokines[148]. Activated HGF (although not total HGF) is increased in cord blood for preterm infants compared to term infants, and HGF increases in the early postnatal period in preterm infants[149]. HGF is also elevated in women with post-partum depression[150]. HGF is involved in alveologenesis, and lower HGF levels are associated with more severe BPD in preterm infants[151], with HGF treatment reducing hyperoxia-induced BPD in a mouse model[152]. | Negative |
| IGFBP4 (Insulin-like growth factor-binding protein 4) | IGFBPs form a complex with IGFs in the circulation, with IGFBP4 inhibiting their effects, and present in serum follicular fluid, seminal fluid, interstitial fluid and synovial fluid[153]. One role of IGFBP4 is related to growth and metabolism[136]. Early pregnancy high IGFBP4 is associated with the development of fetal growth restriction[154]. IGFBP4 is increased by inflammatory cytokines in an animal model of lung injury[155], and is involved in immune responses through IGF1 and the balance between T helper 17 and regulatory T cells[156]. It is also a mediator of acute and chronic stress, with reactions to various genotoxic exposures and mediating aging actions[157]. | Negative |
| L-selectin (SELL, CD62L) | L-selectin is a cell adhesion molecule present on leukocytes, with roles in immune cell adhesion, migration, and signal transduction[158]. It may have a particular role in response to virus infection[158], and is raised on day 7 in preterm infants who develop BPD[159]. It is decreased in neonates with severe hypoxia[160]. L-selectin gene expression is increased in umbilical cord tissue from preterm infants[49], but decreased in the cord blood of preterm neonates after gestational diabetes[107]. L-selectin increases between birth and term corrected GA in preterm infants[20]. | Positive |
| Lactotransferrin (Lactoferrin) | Lactotransferrin is an iron-binding glycoprotein, present in tears, saliva, bile, pancreatic fluid, vaginal secretions, semen, and milk[161]. It supports bacterial and viral immune responses, through direct effects and cytokine stimulation[162]. Lactotransferrin supplementation has been clinically investigated, and reduces rates of late-onset sepsis preterm infants, but does not reduce NEC alone on systematic review[163]. Lactotransferrin has roles in neuroinflammation, with involvement in Alzheimer’s disease and Parkinson’s disease[162], and neuroprotection in animal models of neonatal hypoxic ischaemic encephalopathy and preterm brain injury[162], but thus far clinical supplementation does not appear to improve neurodevelopment[163]. Lactotransferrin also regulates bone formation through osteoblasts[164]. | Positive |
| MMP9 (Matrix metalloproteinase-9) | MMP9 is part of a family of enzymes involved in angiogenesis, cell migration, and invasion, and expression is stimulated by various growth factors and cytokines[165]. It plays a role in brain development through the extracellular matrix[166]. MMP9 is decreased in cord blood[76,167] but increased in blood in the days after birth[77], and in CSF in the first weeks of life[168] in preterm infants compared to term infants. MMP9 gene expression is increased in umbilical cord tissue from preterm infants[49]. It is also increased in vitreous and tear samples of babies with ROP[169]. Concentrations of MMP9 in the first weeks of life are also associated with ventriculomegaly[78], and increased likelihood of executive dysfunction and ADHD symptoms at 10 years of age[79,80], although not cerebral palsy[81]. MMP9 is decreased in the CSF of infants with post-haemorrhagic ventricular dilatation (PHVD)[170]. MMP9 is a urinary biomarker of prematurity-associated lung disease in school-age children[171]. MMP9 EpiScore has been associated with rheumatoid arthritis, lung cancer, cardiovascular disease, and COPD in adults[3,17]. | Negative |
| NCAM1 (Neural cell adhesion molecule 1) | NCAM1 is a synaptic adhesion molecule, part of the immunoglobulin superfamily, involved in neural cell differentiation and migration, neurite outgrowth, synaptic plasticity, and signalling[172]. NCAM1 is decreased in the CSF of infants with PHVD[170], and negatively correlated with neurodevelopment[173]. It has been associated with a range of neuroinflammatory processes such as autism[174], bipolar disorder[175], and schizophrenia[176], and is a mediator of stress structural and functional changes in the brain[177]. NCAM1 is differentially expressed in the placenta with spontaneous miscarriage[178], and preterm birth[179]. NCAM1 also correlates with BMI[137], and raised in the urine of preterm babies with non-infectious respiratory disease compared to infections respiratory disease or control[180]. | Positive |
| PIGR (Polymeric immunoglobulin receptor) | PIGR is the receptor for polymeric IgA and IgM in epithelial cells, leading to secretory IgA and IgM release as part of the immune response to infections, increased by inflammatory cytokines[181,182]. Expression of PIGR and an individual’s microbiome are bidirectionally related[183]. PIGR is also upregulated in children with PDA[184]. | Negative |
| S100A9 (Calgranulin B, MRP14) | S100A9 is a calcium binding protein, expressed in neutrophils and monocytes, stimulating leukocyte recruitment and cytokine release during an inflammatory response, and combining with S100A8 to form calprotectin[185]. S100A9 is involved in bacterial and viral infections, metabolic inflammation such as obesity, autoimmune conditions such as psoriatic arthritis and SLE, and Alzheimer’s disease[185]. S100A9 gene expression is increased in umbilical cord tissue from preterm infants[49], and in maternal serum during pregnancies leading to preterm birth[186]. Calprotectin regulates the development of the microbiome, and is lower in preterm infants[187]. | Positive |
| SCGF alpha (Stem cell growth factor alpha, CLEC11A, C-type lectin domain containing 11a, osteolectin) | SCGFs are positive regulators of cell proliferation, both haematopoietic growth factors, and promoters of osteogenesis[188]. CLEC11A is a urinary biomarker of prematurity-associated lung disease in school-age children[171]. Although a specific immune function for CLEC11A is not currently known, other C-type lectin family members have extensive roles in immunity and homeostasis[189]. SCGF alpha is associated with pulmonary vascular disease in preterm infants[190]. SCGF alpha EpiScore is associated with IBD[3]. | Positive |
| SCGF beta (Stem cell growth factor beta, CLEC11A, C-type lectin domain containing 11a, osteolectin) | SCGFs are positive regulators of cell proliferation, both haematopoietic growth factors, and promoters of osteogenesis[188]. CLEC11A is a urinary biomarker of prematurity-associated lung disease in school-age children[171]. Although a specific immune function for CLEC11A is not currently known, other C-type lectin family members have extensive roles in immunity and homeostasis[189]. SCGF beta is increased in idiopathic inflammatory myopathies[47]. SCGF beta is associated with pulmonary vascular disease in preterm infants[190]. | Positive |
| Semaphorin 3E (SEMA3E) | Semaphorin 3E binds to plexin D1 to modulate the immune response through cell migration and proliferation, and cytokine release, particularly in LPS-induced inflammation[191]. Semaphorin 3E also regulates neuron axonal growth, particularly in the development of the hippocampus[192], and is anti-angiogenic so has been proposed as a therapeutic option for ROP[193,194]. It also has roles in malignancies[195] and immune conditions such as systemic sclerosis, asthma[195], and ulcerative colitis[196]. Semaphorin 3E mutations have also been found in CHARGE syndrome[197]. Semaphorin 3E EpiScore is reduced in COPD[3]. | Positive |
| Sialoadhesin (SIGLEC1, CD169) | Sialoadhesin is a sialic acid-binding protein expressed on leukocytes, with a role stimulating immune responses, such as to bacterial and viral infections[198,199], but also autoimmune conditions such as SLE[200], MS[201], and congenital heart block[202]. | Negative |
| SKR3 (Serine/threonine-protein kinase receptor R3, ACVRL1, Activin receptor-like type 1) | SKR3 is a receptor for transforming growth factor beta (TGF-beta) ligands, with its main role in angiogenesis and vascular regulation. SKR3 mutations lead to hereditary haemorrhagic telangiectasia type 2[203], and it is involved in development of the neonatal lung[204], and in Alzheimer’s disease progression[205]. SKR3 EpiScore is associated with depression and COPD[3]. | Negative |
| SLITRK5 (SLIT and NTRK-like protein 5) | SLITRK5 is a transmembrane protein expressed across the CNS, involved in axon and dendrite growth, neuron differentiation and synaptogenesis[206]. It has been linked to autism, ADHD, obsessive compulsive disorder, and Parkinson’s disease[206]. It is also involved in the regulation of osteoblasts[207], and is downregulated in the placentas from infants with neonatal opioid withdrawal syndrome[208]. | Positive |
| Thrombopoietin receptor (TPOR, MPL, myeloproliferative leukaemia protein) | The thrombopoietin receptor regulates platelet production[209], and low expression in neonates and preterm infants specifically leads to thrombocytosis[210]. Thrombopoietin receptor EpiScore is associated with diabetes[3]. | Positive |
| Trypsin-2 (PRSS2, TRY2, protease serine 2) | Trypsin-2 is produced by the pancreas, and upregulated in pancreatitis[211], and IBD[212]. Trypsin-2 is increased in the tracheal aspirates of infants developing BPD[159]. Low trypsin-2 in pregnancy is predictive of developing pre-eclampsia[213]. Trypsin-2 EpiScore is associated with diabetes, COPD and IHD[3]. | Negative |
| VCAM1 (Vascular cell adhesion protein 1) | VCAM1 is involved in leukocyte-endothelial cell adhesion, playing a role in immune responses through leukocyte migration[214]. In adults, it is involved in various inflammatory diagnoses including rheumatoid arthritis, asthma, transplant rejection, and cancer[215]. VCAM1 concentrations are increased in CSF in preterm infants compared to term infants[216], and in blood concentrations vary by birth GA and chronological age but are not associated with placental inflammation[217]. VCAM1 gene expression is increased in umbilical cord tissue from preterm infants[49]. Lower concentrations of VCAM1 in the first weeks of life are associated with increased likelihood of executive dysfunction at 10 years of age[79], and higher concentrations of VCAM1 with echogenic brain lesions[78] and an increased likelihood of ADHD symptoms[80], although not cerebral palsy[81]. In cord blood, increased VCAM1 is associated with an increased likelihood of autism[104]. VCAM1 EpiScore is associated with cardiovascular disease in adults[17]. | Negative |
| VEGFA (Vascular endothelial growth factor A) | VEGF is a proinflammatory protein involved in angiogenesis and vasculogenesis through endothelial cell proliferation and migration, as well as endothelial permeability, and is stimulated by proinflammatory cytokines[218,219]. VEGF concentrations vary by birth GA and chronological age but are not associated with placental inflammation[217]. VEGFA gene expression is increased in umbilical cord tissue from preterm infants[49]. Raised cord blood VEGF is associated with a reduced likelihood of postnatal growth failure[220]. VEGF is increased in blood, vitreous and tear samples of babies with ROP[15,169], and indeed anti-VEGF monoclonal antibodies are used in the treatment of ROP[221]. Lower serum VEGF is seen in preterm infants with respiratory distress syndrome[222], and VEGF has also been linked to BPD, although primarily in animal studies[223]. Concentrations of VEGFR1 and VEGFR2, but not VEGF, in the first weeks of life are associated with ventriculomegaly[78] and increased likelihood of executive dysfunction at 10 years of age[79], although concentrations of VEGF, VEGFR1 and VEGFR2 are all associated with ADHD symptoms[80]. However, concentrations were not associated with cerebral palsy[81]. VEGFA EpiScore is associated with COPD, cardiovascular disease, and diabetes in adults[3,17], and methylation of the VEGFA gene region changed around the time of NEC diagnosis[224]. | Negative |
| WFIKKN2 (WAP, Kazal, immunoglobulin, Kunitz and NTR domain-containing protein 2, GASP1, Growth and differentiation factor-associated serum protein 1) | WFIKKN2 is involved in changing the presentation of transforming growth factor beta family proteins to their receptors[225], including binding to GDF8 and GDF11[226]. There is a bidirectional relationship between WFIKKN2 and obesity in adults[137], and correlates with BMI[137]. WFIKKN2 decreases between birth and term corrected GA in preterm infants[20]. | Positive |

43 EpiScores associated with birth GA in regression models adjusted for socioeconomic status. Roles adapted from the STRING database[227] with additional references as required.

**Supplementary Figure 10. Interaction between afamin EpiScore, birth gestational age, and maternal education**

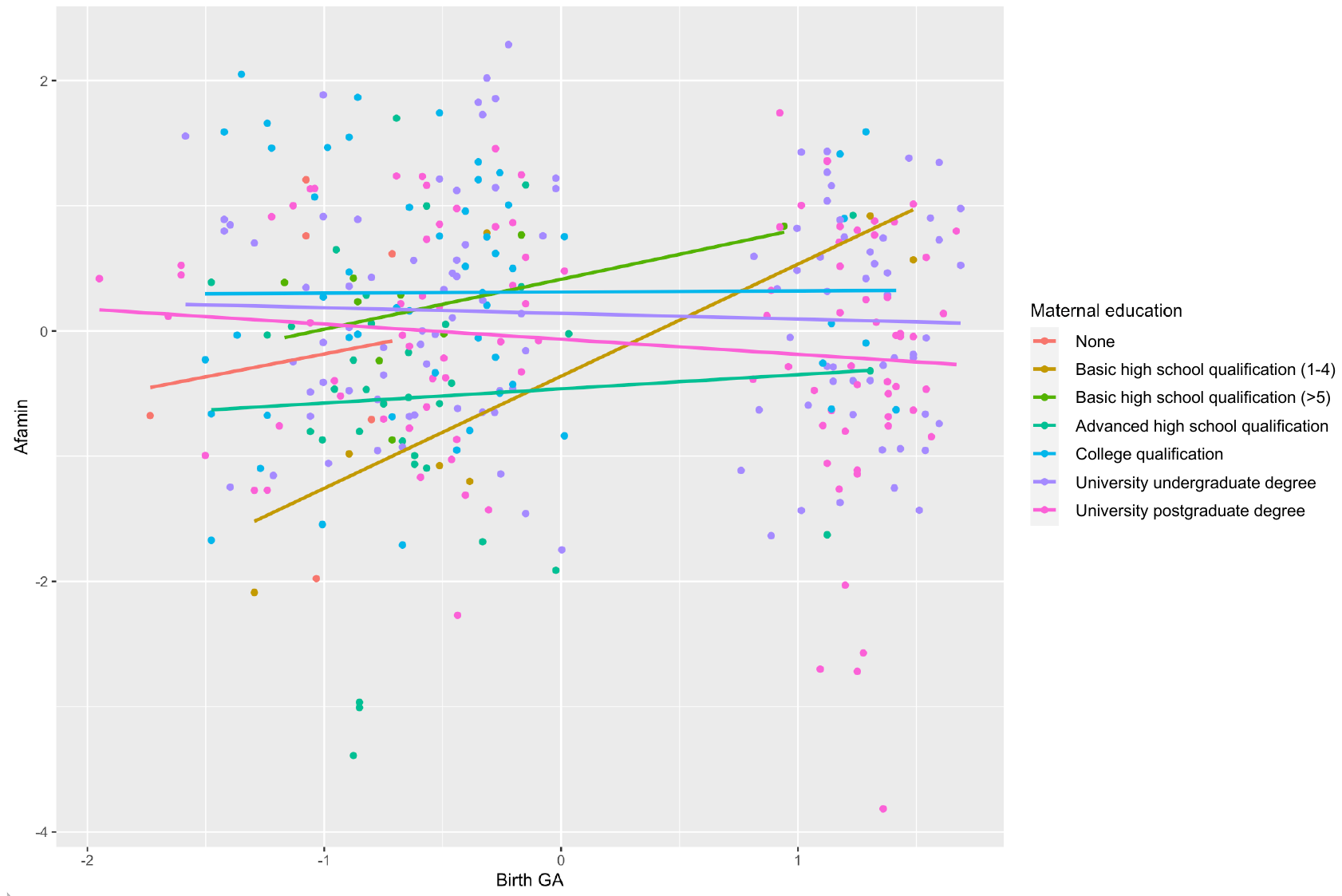

Relationship between Afamin EpiScore and birth gestational age by maternal education. n=323.

GA – gestational age

**Supplementary Figure 11. Interaction between HGFI EpiScore, birth gestational age, and maternal occupation**

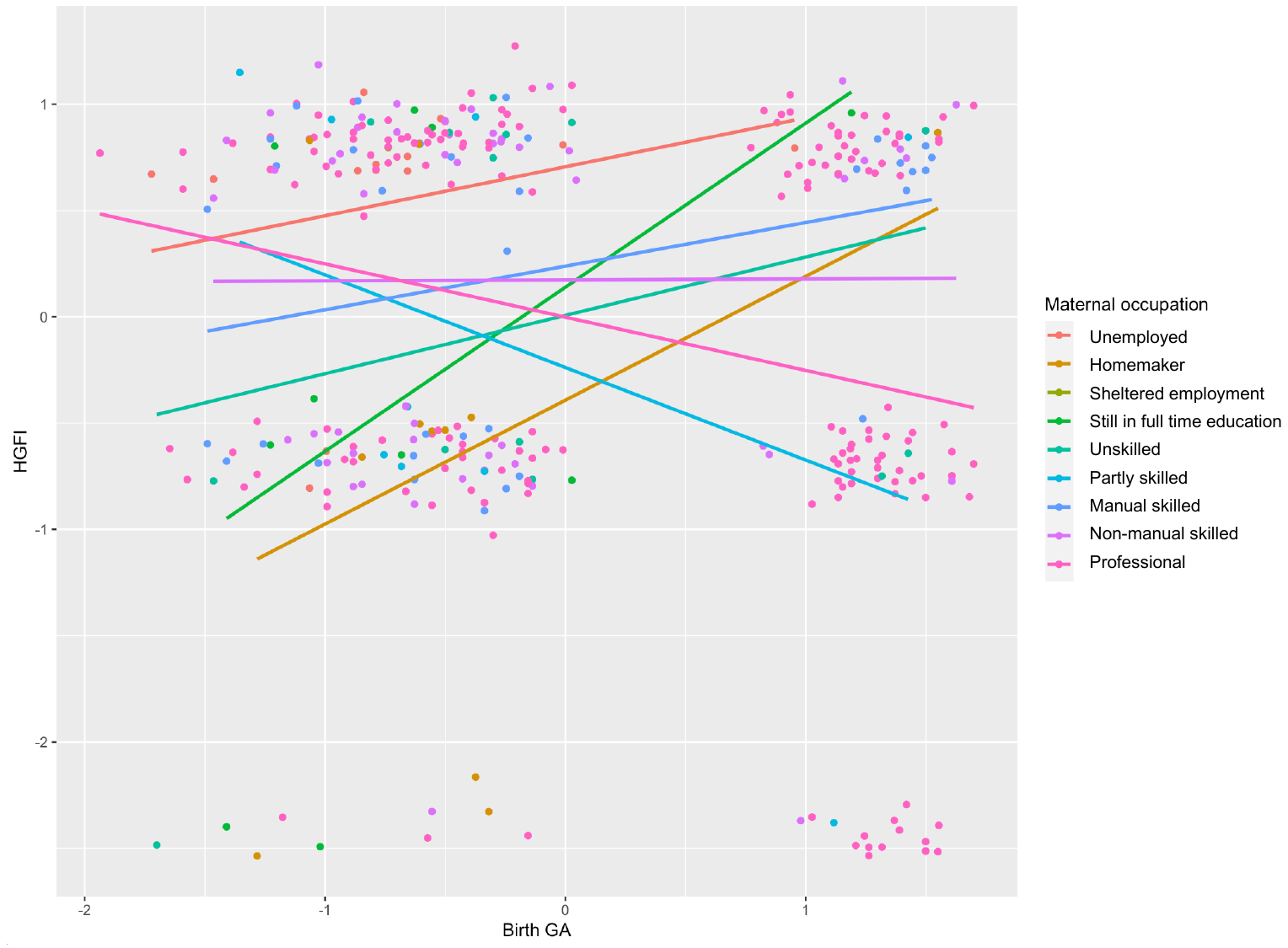

Relationship between HGFI EpiScore and birth gestational age by maternal occupation. n=328.

GA – gestational age, HGFI – hepatocyte growth factor-like protein

**Supplementary Table 5. Relationship between EpiScores and socioeconomic status measure in unadjusted and adjusted models**

| **EpiScore** | **Socioeconomic status measure** | **Unadjusted** | | | **Adjusted** | | |
| --- | --- | --- | --- | --- | --- | --- | --- |
|  |  | **Standardised beta (95% CI)** | ***p-*value** | **R^2^** | **Standardised beta (95% CI)** | ***p-v*alue** | **R^2^** |
| ICAM5 | SIMD | 0.132 (0.035-0.229) | 0.0079 | 0.259 | 0.134 (0.002-0.266) | 0.0454 | 0.369 |
| Afamin | Maternal occupation | 0.061 (0.016-0.107) | 0.0082 | 0.273 | 0.045 (-0.006-0.096) | 0.0827 | 0.432 |
| Afamin | Birth GA* maternal education | -0.11 (-0.198- -0.038) | 0.0041 | 0.278 | 0.014 (-0.082-0.110) | 0.7745 | 0.437 |
| HGFI | Birth GA*  maternal occupation | 0.097 (-0.159- -0.036) | 0.0021 | 0.053 | -0.013 (-0.077-0.051) | 0.6810 | 0.029 |

EpiScores associated with socioeconomic status (Scottish Index of Multiple Deprivation, maternal education, or maternal occupation), or with an interaction between socioeconomic status and birth gestational age. Unadjusted regression models are for all included infants (n=332), and include GA at sample, birthweight z-score, sex, and methylation processing batch. Adjusted regression models are for preterm infants only (n=115), and additionally include inflammatory exposures; sepsis, histological chorioamnionitis, necrotising enterocolitis, and bronchopulmonary dysplasia. Adjusted *p-*value <8.3x10^-3^.

CI – confidence interval, GA – gestational age, HGFI – Hepatocyte growth factor-like protein alpha chain, ICAM5 – Intercellular adhesion molecule 5, SIMD – Scottish Index of Multiple Deprivation.
